## Supplementary Tables/Figures for "Nanopore Metagenomic Sequencing for Detection and Characterization of SARS-CoV-2 in Clinical Samples"

**Supplementary Material**

**Supplementary Table 1: A.** Primers and **B.** probes for RT-PCR testing performed at the BCCDC Public Health Laboratory and VGH

| A. Primer Name | Sequence (5′ → 3′) | Target | Design |
|---|---|---|---|
| **BCCDC RdRP** | | | |
| BCCDC RdRP Fwd | TGCCGATAAGTATGTCCGCA | RNA-dependent RNA polymerase, SARS-CoV-2 specific | Tracy Lee, BCCDC |
| BCCDC RdRP Rev | CAGCATCGTCAGAGAGTATCATCATT | | |
| **E-gene** | | | |
| E-gene_Fwd | ACAGGTACGTTAATAGTTAATAGCGT | Envelope protein, all SARS related viruses | Tracy Lee, BCCDC |
| E-gene_Rev | ATATTGCAGCAGTACGCACACA | | |
| **RNAseP** | | | |
| RNAseP F | AGATTTGGACCTGCGAGCG | Human ribonuclease P | Tracy Lee, BCCDC |
| RNAseP R | GAGCGGCTGTCTCAACAAGT | | |

| B. Probe Names | Sequence (5′ → 3′) |
|---|---|
| **RdRP gene** | |
| BCCDC RdRP probe | ***FAM***/TTGACACAGACTTTGTGAATG/*MGBNFQ* |
| **E-gene** | |
| E-gene _probe | ***Cy5***/ACACTAGCC/*TAO*/ATCCTTACTGCGCTTCG/IAbRQSp |
| **RNAseP** | |
| RNAseP probe | ***NED***/TCTGACCTGAAGGCTC/*MGBNFQ* |

Mixed bases: **Y** = C or T, **K** = G or T, **R** = A or G

**Supplementary Table 2: A.** Primers, **B.** probes, and **C.** reaction conditions for Variant of Concern PCR testing

| **A. Primers** | | | |
|---|---|---|---|
| **Primer name** | **target** | **Sequence 5'-3'** | **Design** |
| N501Y_E484K-F1 | E484K | AGAGAGATATTTCAACTGAAATCTATCAGG | Tracy Lee, BCCDC |
| N501Y-R3 | E484K & N501Y | CCACAAACAGTTGCTGGTGC | Tracy Lee, BCCDC |
| N501Y-F2 | N501Y | AATTGTTACTTTCCTTTACAATCATATGG | Tracy Lee, BCCDC |
| WT_N501Y_block | N501Y | CCAAC+C+CA+C+T+A+A/3InvdT/ | Tracy Lee, BCCDC |
| E_Sarbeco_F1 | E-gene | ACAGGTACGTTAATAGTTAATAGCGT | Corman et al. |
| E_Sarbeco_R2 | E gene | ATATTGCAGCAGTACGCACACA | Corman et al. |

| **B. Probes** | | | |
|---|---|---|---|
| **Target** | **Probe Dye** | **Sequence 5'-3'** | **Design** |
| E484K_MGB-P1 VIC | VIC-MGB | CTTGTAATGGTGTTAAAGGT | Tracy Lee |
| N501Y-P_FAM MGB | FAM-MGB | CCAACCCACTTATGG | Tracy Lee |
| E_Sarbeco_probe | CY5-TAO | ACACTAGCCATCCTTACTGCGCTTCG | Corman et al. |

**C. Thermocycling program**

| **Step** | **Cycles** | **Temperature** | **Time** |
|---|---|---|---|
| RT | 1 | $50^0$C | 5 minutes |
| Enzyme Activation | 1 | $95^0$C | 20 seconds |
| Amplification | 40 | $95^0$C | 3 seconds |
| | | $60^0$C | 30 seconds |

**Supplementary Table 3:** Detection of potential pathogens in study samples.

| Study ID | Bacterial Reads | Co-infection (% total bacterial/viral reads) |
|---|---|---|
| P1 | 9,553 | *Moraxella Catarrhalis* (7.4%) |
| P2 | 64,987 | *Haemophilus influenzae* (2.8%) |
| P4 | 24,479 | *Haemophilus parainfluenzae* (16.8%), *Neisseria meningitidis* (9.6%) |
| P7 | 3,896 | *Haemophilus influezae* (14.7%), *Haemophilus parainfluenzae* (6.9%) |
| P8 | 171,862 | Haemophilus parainfluenzae (1.6%) |
| P9 | 3,132,744 | *Haemophilus parainfluenzae* (1.1%) |
| P13 | 619,910 | *Haemophilus parainfluenzae* (4.5%), *Streptococcus pneumoniae* (2.7%) |
| P14 | 481,963 | *Moraxella catarrhalis* (31.6%) |
| P17 | 307,454 | *Haemophilus parainfluenzae* (3.5%) |
| P22 | 2,121 | *Neisseria meningitidis* (4.6%), *Klebsiella pneumoniae* (1.1%) |
| P27 | 2,862 | *Klebsiella pneumoniae* (69.3%) |
| P36 | 829 | *Staphylococcus aureus* (9.5%) |
| P38 | 7,376 | *Staphylococcus aureus* (10.1%) |
| N1 | 1,192 | *Staphylococcus aureus* (54.4%) |
| N5 | 55,465 | *Streptococcus pneumoniae* (59.5%) |

Only pathogens with greater than 1% relative abundance in samples with at least 1000 bacterial reads were included.

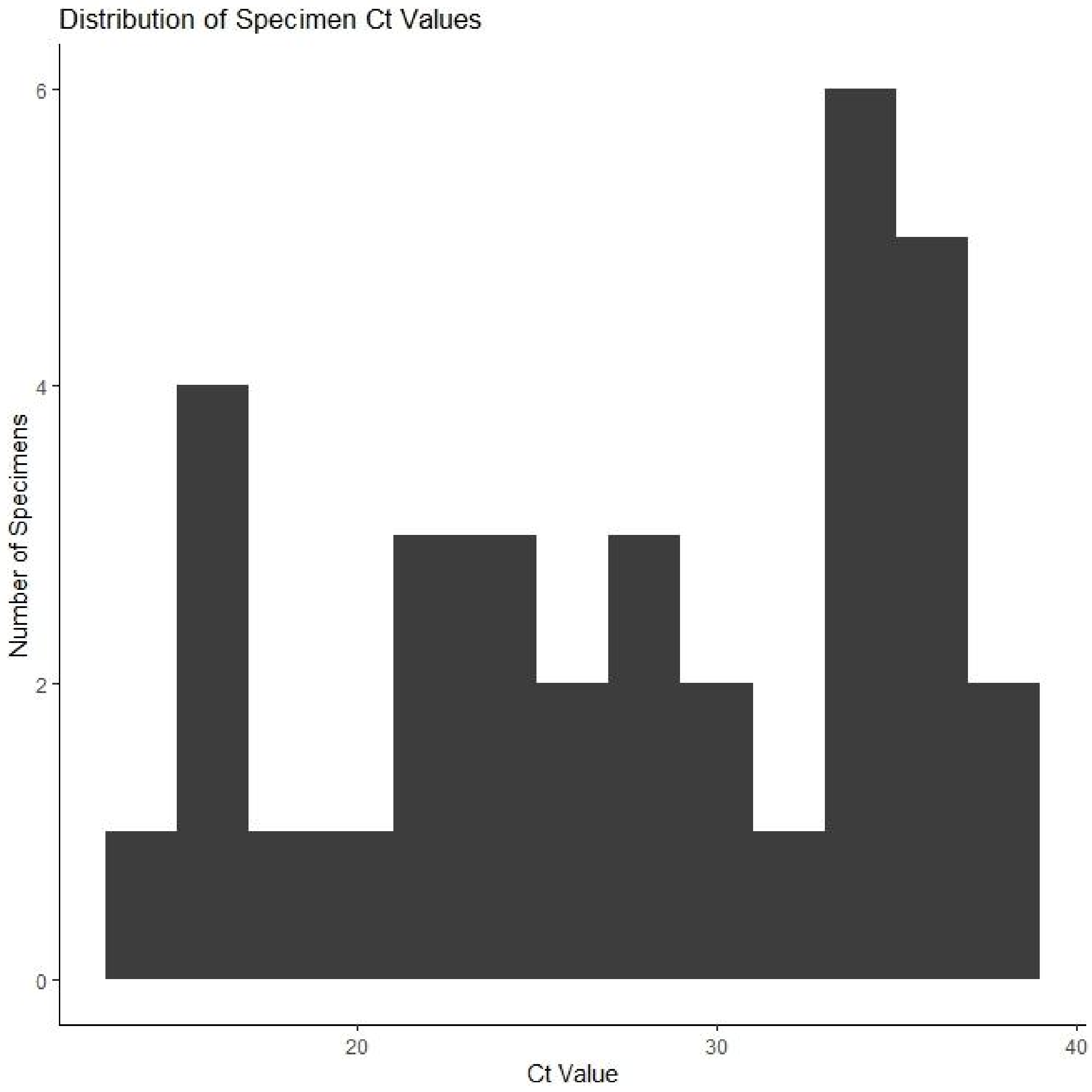


**Supplementary Figure 1:** Distribution of SARS-CoV-2 RT-qPCR positive study samples across $C_t$ values

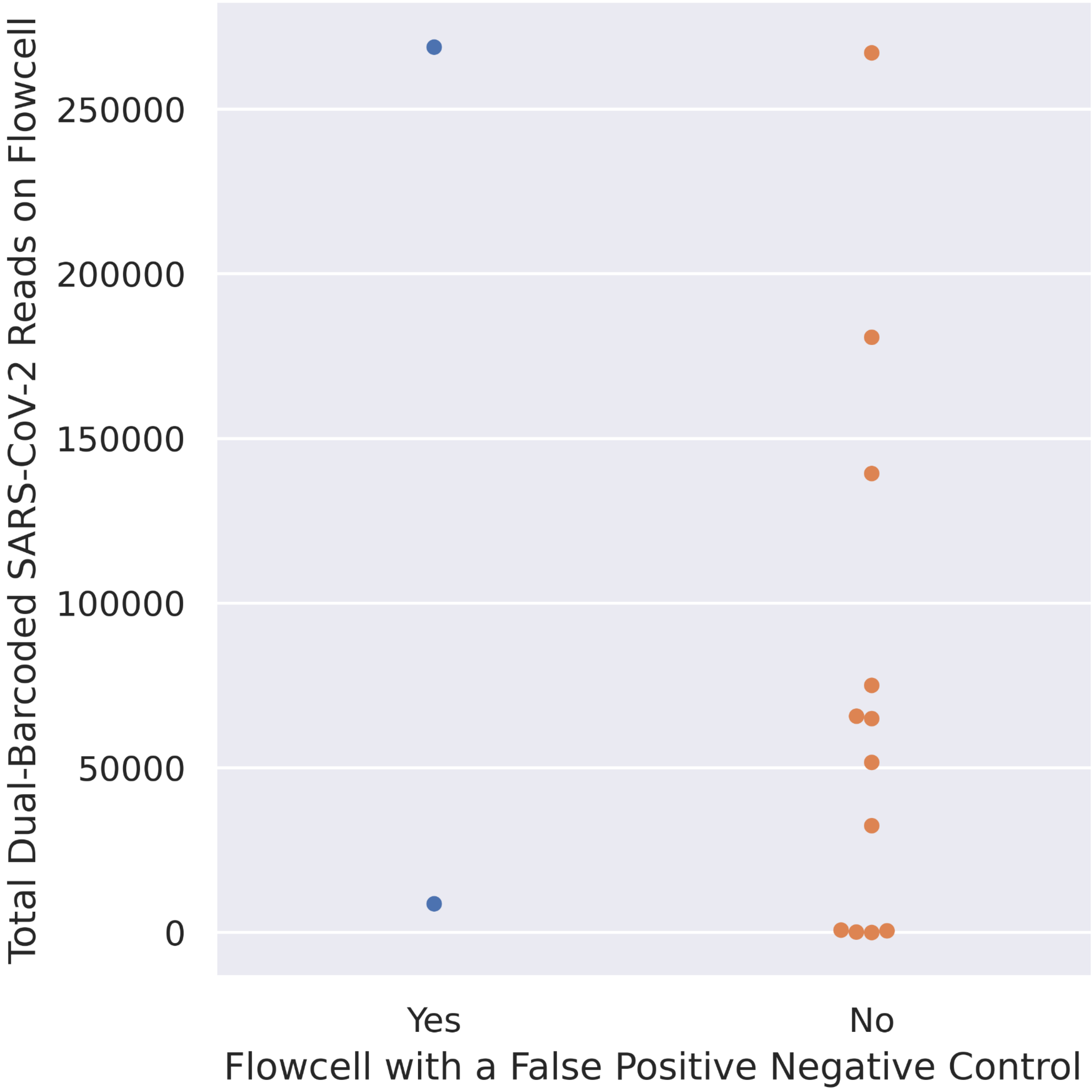


**Supplementary Figure 2:** Comparison of the total number of SARS-CoV-2 reads across all samples on a flowcell, stratified by flowcells with and without a false-positive negative control.

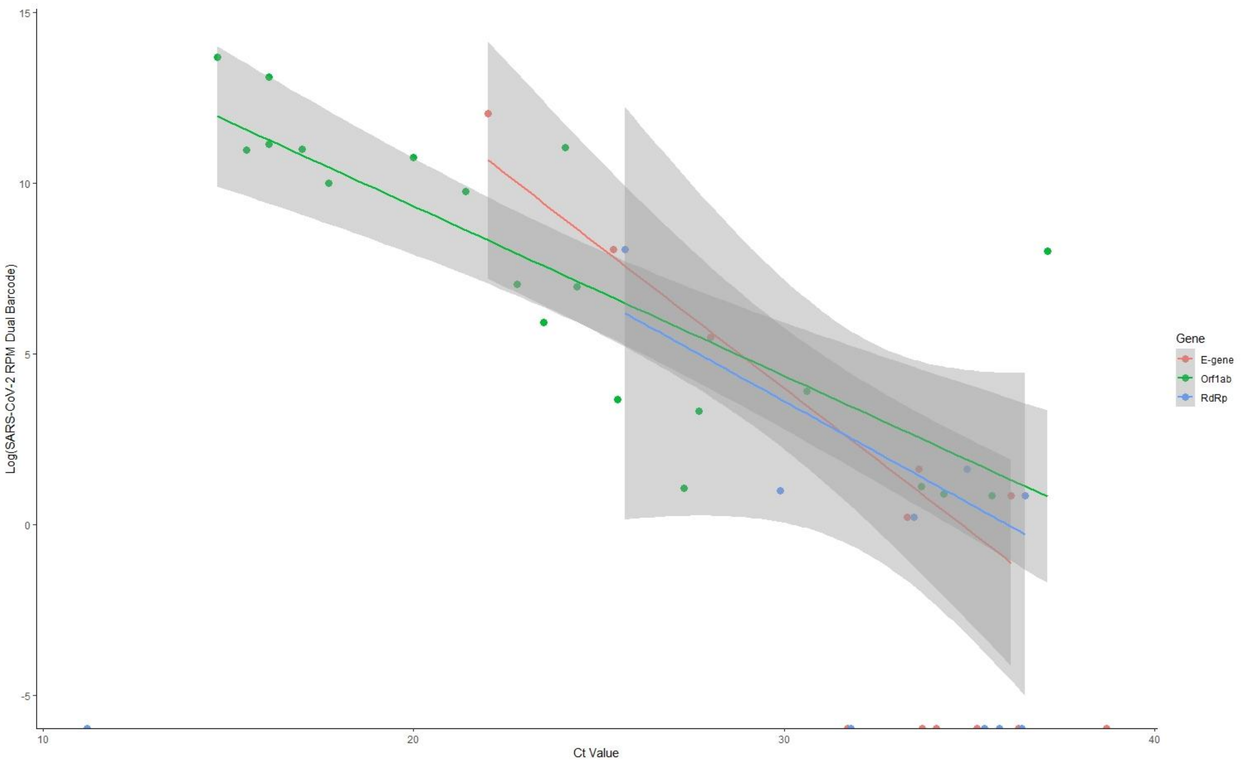


**Supplementary Figure 3:** Scatterplot of Log(SARS-CoV-2 RPM) against $C_t$ value stratified by E-gene, ORF1ab, or RdRp.

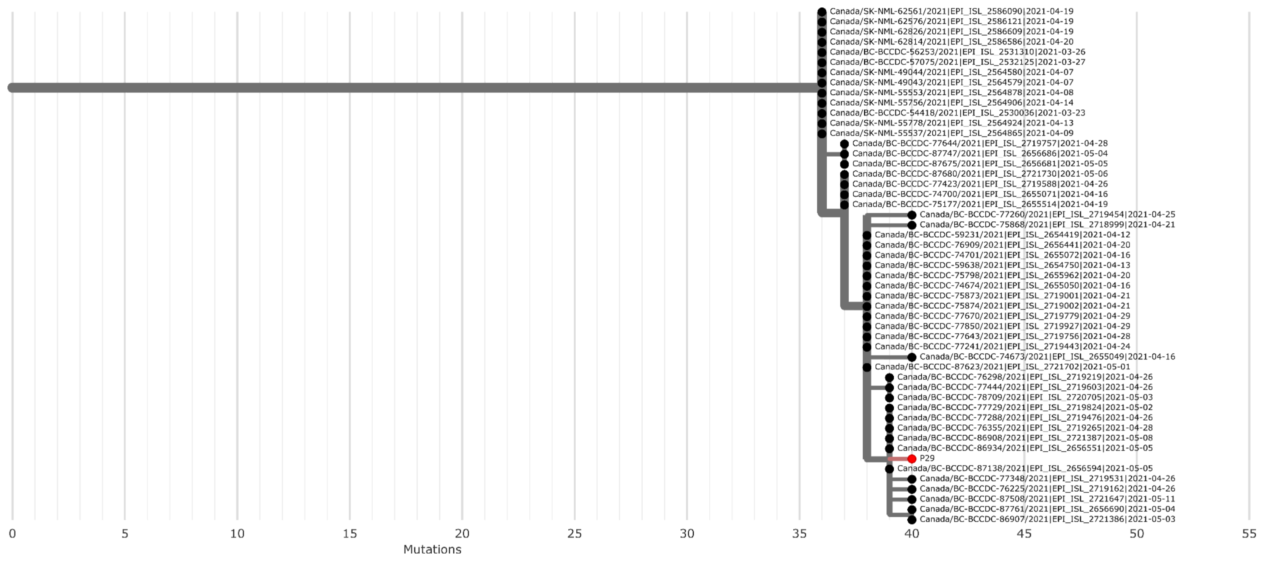
P29
0
5
10
15
20
25
30
35
40
45
50
55
Mutations

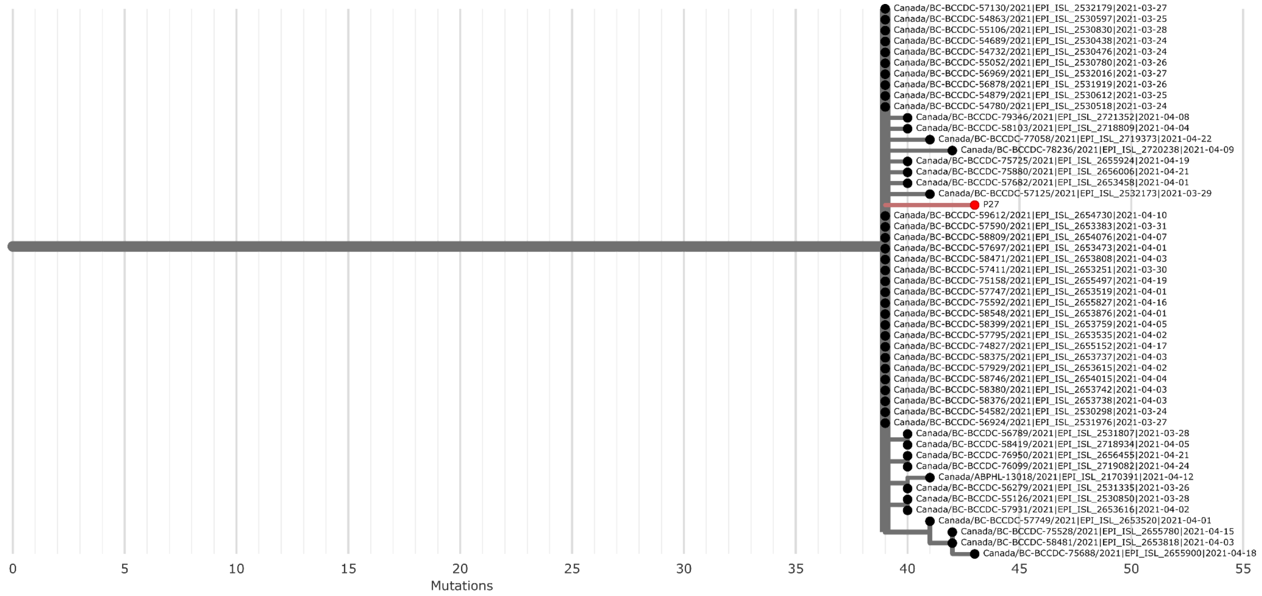
P27
0
5
10
15
20
25
30
35
40
45
50
55
Mutations

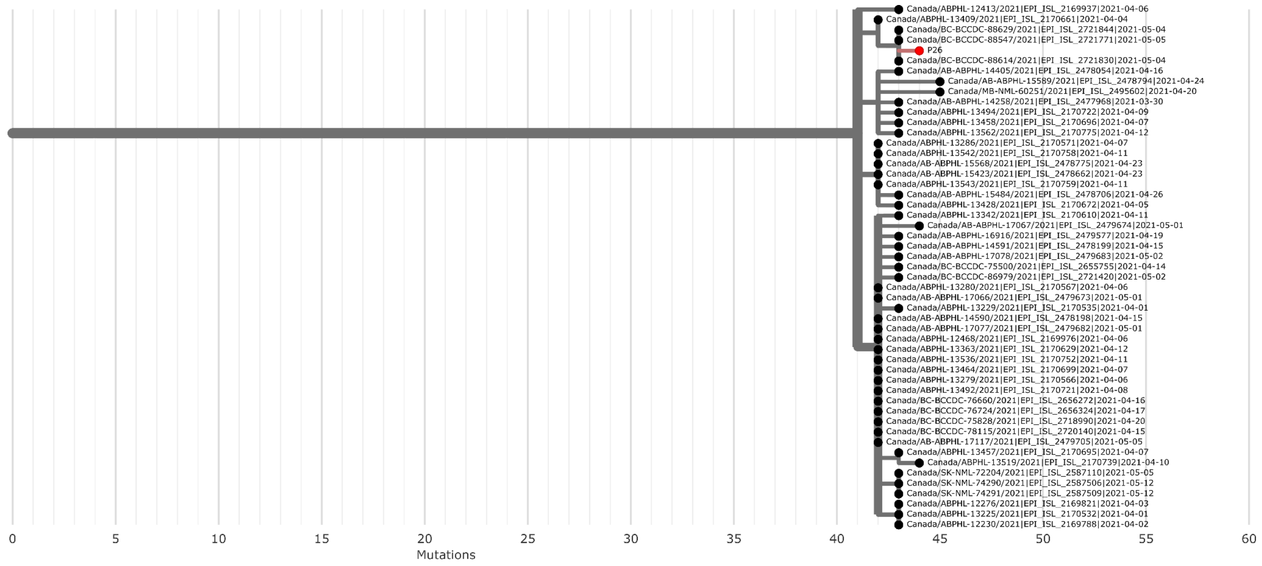
P26
0
5
10
15
20
25
30
35
40
45
50
55
60
Mutations

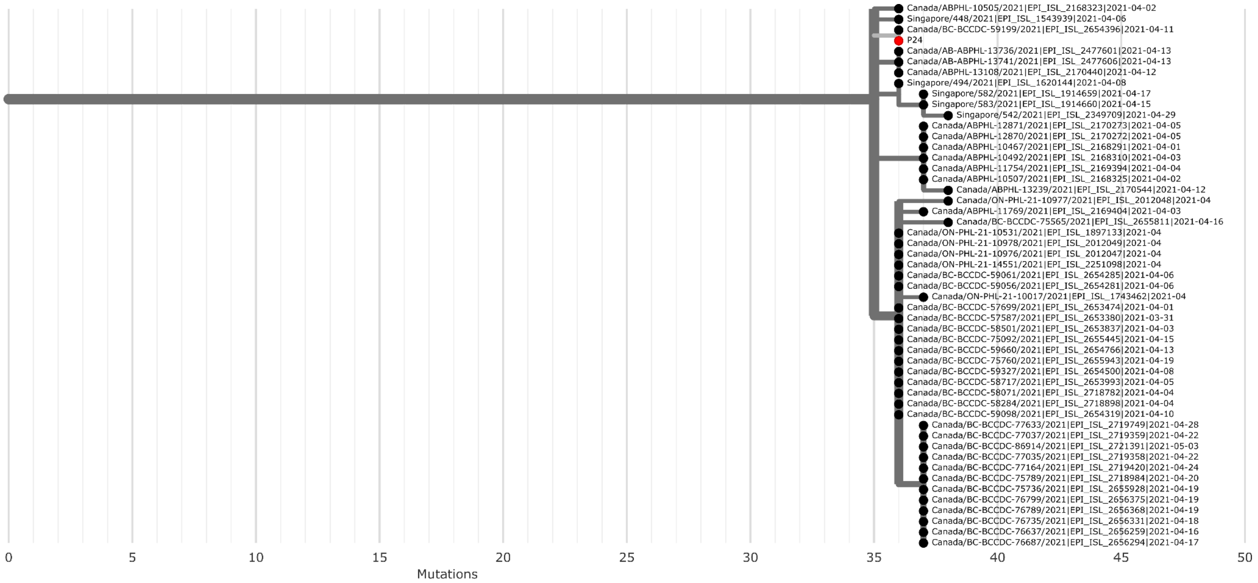
P24
0
5
10
15
20
25
30
35
40
45
50
Mutations

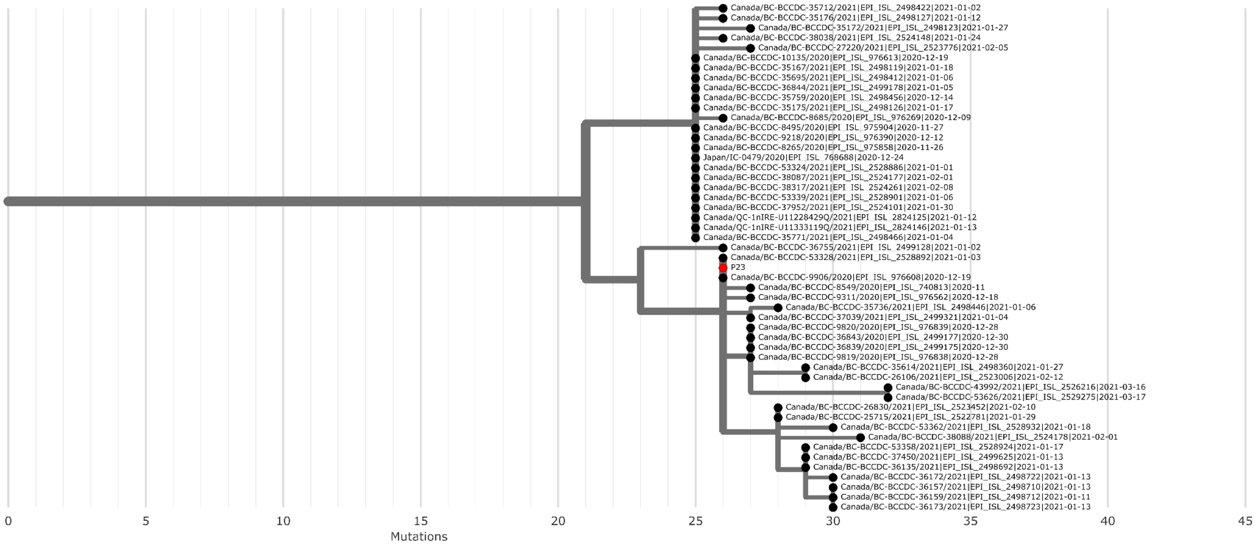
P23
0
5
10
15
20
25
30
35
40
45
Mutations

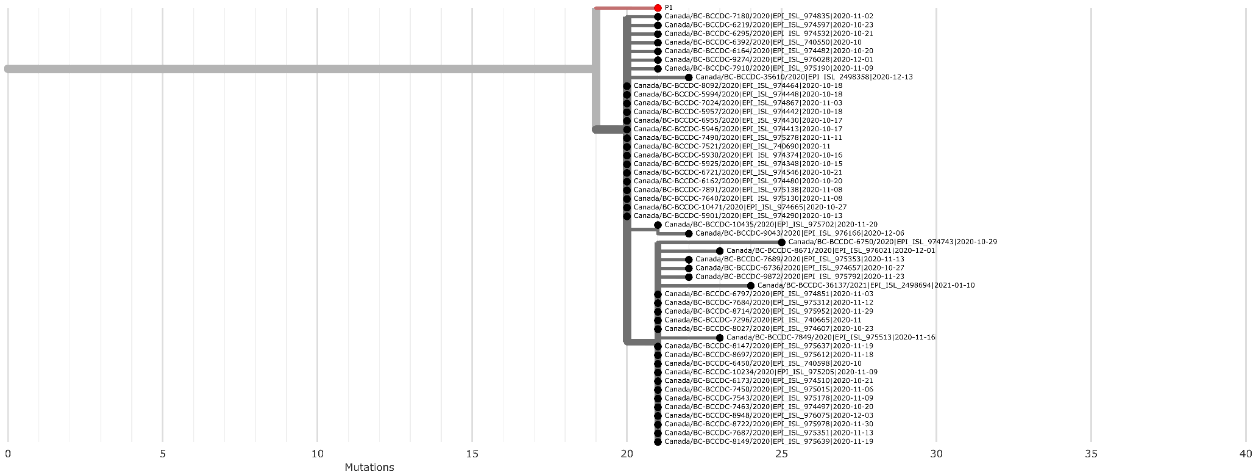
P1
0
5
10
15
20
25
30
35
40
Mutations

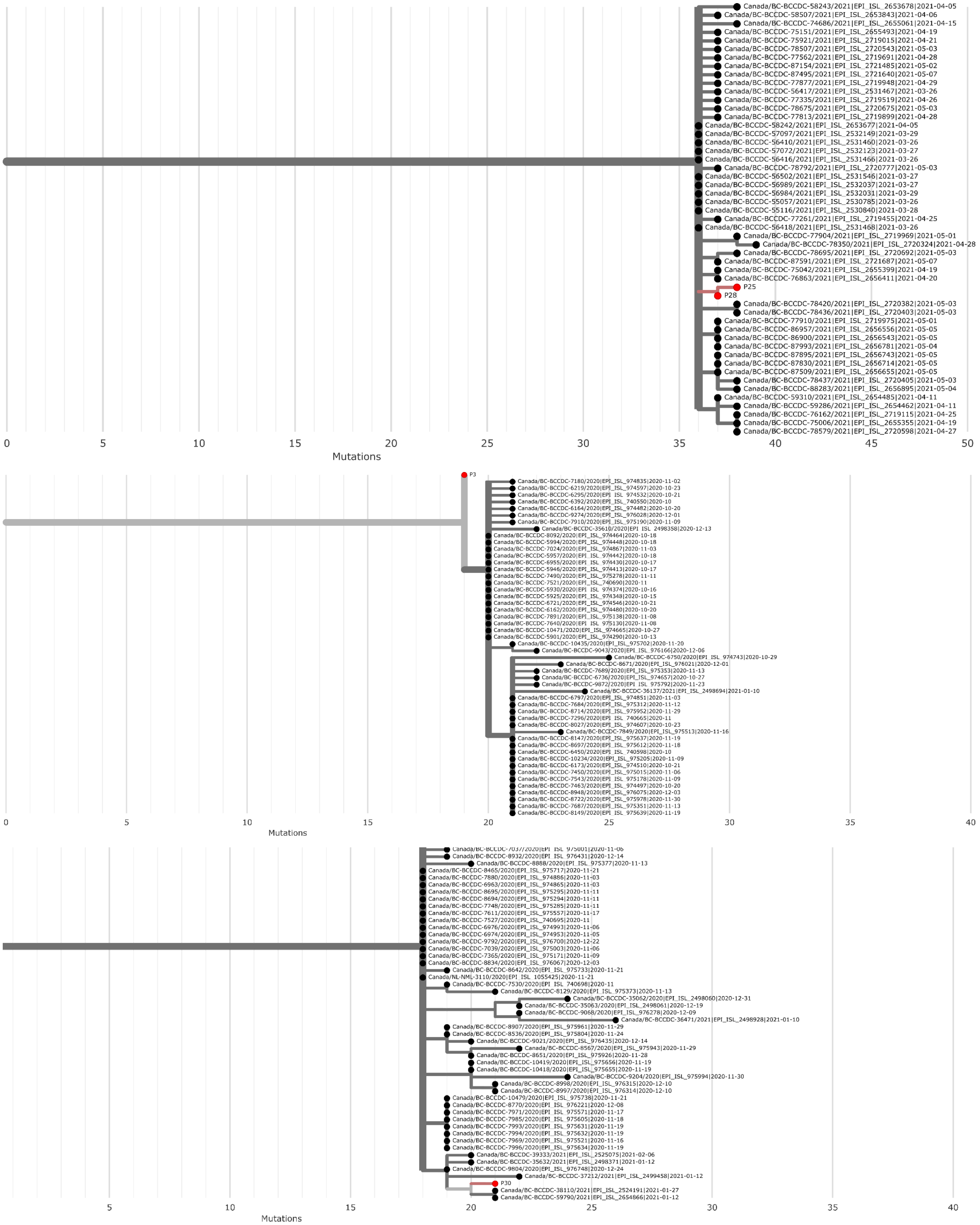


**Supplementary Figure 4:** Phylogenetic Subtrees of Study Samples Compared with 2,447,008 publicly available SARS-CoV-2 genomes. Study samples are marked in red, publicly available genomes are black.

**Supplementary Data:** Representative visualization file for BugSeq metagenomic classification results produced by Recentrifuge
