## Supplementary Data for "Nanopore Metagenomic Sequencing for Detection and Characterization of SARS-CoV-2 in Clinical Samples"

 Javascript must be enabled to view this page.

countunassignedtidrankscore0c8ccd2b\_barcode860c8ccd2b\_barcode870c8ccd2b\_barcode880c8ccd2b\_barcode890c8ccd2b\_barcode900c8ccd2b\_noBarcodestringent\_demultiplex\_0c8ccd2b\_barcode86stringent\_demultiplex\_0c8ccd2b\_barcode87stringent\_demultiplex\_0c8ccd2b\_barcode88stringent\_demultiplex\_0c8ccd2b\_barcode89stringent\_demultiplex\_0c8ccd2b\_barcode90stringent\_demultiplex\_0c8ccd2b\_noBarcode0c8ccd2b\_barcode86\_EXCLUSIVE\_species0c8ccd2b\_barcode87\_EXCLUSIVE\_species0c8ccd2b\_barcode88\_EXCLUSIVE\_species0c8ccd2b\_barcode89\_EXCLUSIVE\_species0c8ccd2b\_barcode90\_EXCLUSIVE\_species0c8ccd2b\_noBarcode\_EXCLUSIVE\_speciesSHARED\_species0c8ccd2b\_barcode86\_EXCLUSIVE\_genus0c8ccd2b\_barcode87\_EXCLUSIVE\_genus0c8ccd2b\_barcode88\_EXCLUSIVE\_genus0c8ccd2b\_barcode89\_EXCLUSIVE\_genus0c8ccd2b\_noBarcode\_EXCLUSIVE\_genusSHARED\_genus0c8ccd2b\_barcode86\_EXCLUSIVE\_family0c8ccd2b\_barcode87\_EXCLUSIVE\_family0c8ccd2b\_barcode88\_EXCLUSIVE\_family0c8ccd2b\_barcode89\_EXCLUSIVE\_family0c8ccd2b\_noBarcode\_EXCLUSIVE\_familySHARED\_family0c8ccd2b\_barcode86\_EXCLUSIVE\_order0c8ccd2b\_barcode87\_EXCLUSIVE\_order0c8ccd2b\_barcode88\_EXCLUSIVE\_order0c8ccd2b\_barcode89\_EXCLUSIVE\_orderSHARED\_order0c8ccd2b\_barcode86\_EXCLUSIVE\_class0c8ccd2b\_barcode89\_EXCLUSIVE\_classSHARED\_class0c8ccd2b\_barcode86\_EXCLUSIVE\_phylumSHARED\_phylum0c8ccd2b\_barcode86\_EXCLUSIVE\_SUMMARY0c8ccd2b\_barcode87\_EXCLUSIVE\_SUMMARY0c8ccd2b\_barcode88\_EXCLUSIVE\_SUMMARY0c8ccd2b\_barcode89\_EXCLUSIVE\_SUMMARY0c8ccd2b\_barcode90\_EXCLUSIVE\_SUMMARY0c8ccd2b\_noBarcode\_EXCLUSIVE\_SUMMARYSHARED\_SUMMARY 317349815973761103117332516228358812468762323983728301010420898354631144261937112922101115137103671376215431344114454526193711445350455307312258116004156353575768752914203611066302191no\_rank71.170.369.169.931.274.169.869.068.069.117.974.463.048.449.156.068.024.077.663.154.250.553.624.076.548.649.345.341.324.076.437.543.07.035.274.772.045.774.146.074.063.148.449.156.068.024.074.038986112120375252349138215821249837251231129189710377333373213431244144538251231144512142122superkingdom55.768.551.766.765.268.356.169.148.667.679.068.368.050.250.558.968.0077.669.359.757.760.8076.560.449.350.747.7076.454.043.07.034.774.772.048.574.1074.068.050.250.558.968.0074.023112091470283229713101826124319143183231132124319111224phylum50.752.449.354.852.057.548.246.142.558.1057.368.158.832.759.368.00065.061.79.060.80062.360.5047.70054.059.0034.70048.500068.158.832.759.368.00082487136235943117235451112213211511122321236class45.550.552.353.8054.232.552.745.555.7053.863.664.080.072.800061.064.0082.70061.00089.00054.000000000063.664.080.072.80005143124012855212811191111972274order51.052.261.558.0062.90061.060.5062.972.064.080.069.6000064.00000000000000000000072.064.080.069.60003133117985528851212468family38.751.361.552.0056.20061.046.1056.20080.076.500000000000000000000000000080.076.500031329179754287522469genus38.751.360.952.0057.00061.246.1057.000076.5000000000000000000000000000076.5000122122108980species077.000067.00000067.000000000000000000000000000000000000011111196816no\_rank00079.0092.00000092.000079.0000000000000000000000000000079.00001111112601122species00079.00000000000079.0000000000000000000000000000079.000011112725684species0000092.00000092.00000000000000000000000000000000000001111756892species0000068.00000068.0000000000000000000000000000000000000312291777142871909768species\_group38.749.260.951.7056.00061.246.1056.0000000000000000000000000000000000000312291777142871312291777142871470species38.749.260.951.7056.00061.246.1056.00000000000000000000000000000000000001111112662362species00074.00000000000074.0000000000000000000000000000074.000066475genus0000044.70000044.7000000000000000000000000000000000000111134061species0000084.00000084.0000000000000000000000000000000000000555534062species0000036.80000036.8000000000000000000000000000000000000241411497genus0070.00059.20060.00059.20080.0000000000000000000000000000080.000001441111196806no\_rank0080.00059.20000059.20080.0000000000000000000000000000080.0000011111699623species0000066.00000066.00000000000000000000000000000000000001111111699624species0080.00000000000080.0000000000000000000000000000080.0000011111720344species0000078.00000078.000000000000000000000000000000000000011112203895species0000061.00000061.00000000000000000000000000000000000001111261164species0060.00000060.000000000000000000000000000000000000000021614324431171117135621family69.564.0075.7076.100077.3076.172.064.0067.6000064.00000000000000000000072.064.0067.600026143244317176232286genus69.50075.7076.100077.3076.172.00067.6000000000000000000000000072.00067.60002222104087species0000075.00000075.00000000000000000000000000000000000006422136841species\_group00074.30000078.80000065.5000000000000000000000000000065.500022222287species00065.50000000000065.5000000000000000000000000000065.5000221340851strain00065.500000000000000000000000000000000000000000000111143263species00075.00000075.00000000000000000000000000000000000000022627141species\_subgroup00087.50000087.500000000000000000000000000000000000000222246680species00087.50000087.500000000000000000000000000000000000000111232139species\_subgroup00065.00000065.0000000000000000000000000000000000000001111301species00065.00000065.000000000000000000000000000000000000000111136842species\_group00051.00000000000051.0000000000000000000000000000051.000011111587753species00051.00000000000051.0000000000000000000000000000051.000011587851subspecies00051.0000000000000000000000000000000000000000000001711411111136843species\_group00076.9080.100080.5080.100000000000000000000000000000000000042224222294species00077.5083.500081.0083.5000000000000000000000000000000000000111147879species0000073.00000073.000000000000000000000000000000000000024424447883species00068.0086.80000086.8000000000000000000000000000000000000212175588species00074.00000075.0000000000000000000000000000000000000007212721276758species00078.3074.000085.0074.0000000000000000000000000000000000000111111200451species00077.0066.00000066.000000000000000000000000000000000000011111136845species\_group00070.005.0000005.000070.0000000000000000000000000000070.0000111147880species000005.0000005.000000000000000000000000000000000000011111176759species00070.00000000000070.0000000000000000000000000000070.000014323136846species\_group67.00084.2075.700082.5075.700000000000000000000000000000000000014323578833species\_subgroup67.00084.2075.700082.5075.70000000000000000000000000000000000001432314323316species67.00084.2075.700082.5075.700000000000000000000000000000000000011319619121222196821no\_rank72.00071.8078.900074.2078.972.00066.0000000000000000000000000072.00066.00003333658642species0000084.30000084.300000000000000000000000000000000000011111173270species0000083.00000083.000000000000000000000000000000000000011112018067species0000085.00000085.00000000000000000000000000000000000001111112054914species00078.00000000000078.0000000000000000000000000000078.00001111112498848species72.00000000000072.0000000000000000000000000000072.000000011112604941species0000065.00000065.0000000000000000000000000000000000000343434342653853species00072.7080.500072.7080.500000000000000000000000000000000000022222654238species0000083.50000083.50000000000000000000000000000000000001111112681983species00054.00000000000054.0000000000000000000000000000054.000011112725477species0000078.00000078.000000000000000000000000000000000000011112730846species0000086.00000086.00000000000000000000000000000000000002442442730849species00075.5071.80000071.8000000000000000000000000000000000000411141112730850species00072.5073.000085.0073.00000000000000000000000000000000000001111198618species0000082.00000082.00000000000000000000000000000000000001235312353219572species00077.6083.700074.6083.70000000000000000000000000000000000001111312306species000009.0000009.000000000000000000000000000000000000011111930166species00089.00000000000089.0000000000000000000000000000089.0000186264subspecies00089.00000000000000000000000000000000000000000000011994484strain00089.000000000000000000000000000000000000000000000111111849530genus064.00000000000064.000000064.000000000000000000000064.0000001111111697053species064.00000000000064.0000000000000000000000000000064.0000002229281232212112291347order70.062.074.583.0081.000082.0080.170.00091.000068.00091.00068.00089.000000000000070.00091.000012815114111543family056.074.582.2082.400082.0082.000093.000000093.000000000000000000000093.000011111547genus00093.00000000000093.000000093.000000000000000000000093.0000111354276species\_group00093.00000000000093.0000000000000000000000000000093.0000111111208224species00093.00000000000093.0000000000000000000000000000093.000012715114561genus056.074.580.7082.400082.0082.00000000000000000000000000000000000001271511412715114562species056.074.580.7082.400082.0082.000000000000000000000000000000000000011139111903409family72.068.000079.30000077.172.0000000000000000000000000000072.0000000111391153335genus72.068.000079.30000077.172.0000000000000000000000000000072.00000001111470933species0000076.00000076.00000000000000000000000000000000000002222470934species0000056.50000056.50000000000000000000000000000000000001111111076550species72.00000000000072.0000000000000000000000000000072.0000000221654067species\_group0000084.50000084.50000000000000000000000000000000000002222549species0000084.50000084.50000000000000000000000000000000000001842630326no\_rank068.000084.10000084.00000000000000000000000000000000000001841841484158species068.000084.10000084.00000000000000000000000000000000000001111111903410family00089.00000000000089.000000089.00000089.000000000000000089.000011111122277genus00089.00000000000089.000000089.000000000000000000000089.00001111112108399species00089.00000000000089.0000000000000000000000000000089.00001111111903412family68.00000000000068.000000068.00000068.000000000000000068.000000011111635genus68.00000000000068.000000068.000000000000000000000068.000000011111167780species68.00000000000068.0000000000000000000000000000068.00000001111111118969order54.00000000000054.000000054.00000054.00000054.000000000054.0000000111111118968family54.00000000000054.000000054.00000054.000000000000000054.000000011111776genus54.00000000000054.000000054.000000000000000000000054.00000001112676648no\_rank54.00000000000054.0000000000000000000000000000054.00000001111112487929species54.00000000000054.0000000000000000000000000000054.0000000681811135614order59.8000066.066.0000066.052.0000000000000000000000000000052.000000068181132033family59.8000066.066.0000066.052.0000000000000000000000000000052.0000000681811168genus59.8000066.066.0000066.052.0000000000000000000000000000052.000000012212269species65.0000056.50000056.5000000000000000000000000000000000000313184531species54.30000066.0000000000000000000000000000000000000000001111435897species0000073.00000073.0000000000000000000000000000000000000111111453783species52.00000000000052.0000000000000000000000000000052.0000000552635362no\_rank0000068.40000068.400000000000000000000000000000000000022222290922species0000057.00000057.000000000000000000000000000000000000022222763317species0000075.50000075.500000000000000000000000000000000000011112781024species0000077.00000077.000000000000000000000000000000000000066233510718731416187135619order42.941.742.941.3043.221.37.024.036.8043.20000000000000000000000000000000000006623351071873141618728256family42.941.742.941.3043.221.37.024.036.8043.2000000000000000000000000000000000000662335107187314161872745genus42.941.742.941.3043.221.37.024.036.8043.2000000000000000000000000000000000000662335107187314161872609666no\_rank42.941.742.941.3043.221.37.024.036.8043.200000000000000000000000000000000000066233510718731416187662335107187314161872306583species42.941.742.941.3043.221.37.024.036.8043.2000000000000000000000000000000000000968238111135625order067.8062.7067.0075.5064.3067.000066.000000066.000000000000000000000066.0000968238111712family067.8062.7067.0075.5064.3067.000066.000000066.000000000000000000000066.0000946226724genus067.8061.8071.2075.5065.0071.20000000000000000000000000000000000002131321313726species073.5056.0073.3076.000073.300000000000000000000000000000000000021112111727species00054.5075.000048.0075.0000000000000000000000000000000000000712112712112729species066.1082.0066.0075.0082.0066.000000000000000000000000000000000000011745genus0000061.00000061.00000000000000000000000000000000000001111747species0000061.00000061.000000000000000000000000000000000000011292486genus0000048.00000048.00000000000000000000000000000000000001111762species0000048.00000048.000000000000000000000000000000000000011111416916genus00066.00000000000066.000000066.000000000000000000000066.0000111111739species00066.00000000000066.0000000000000000000000000000066.000011476528genus00063.00000063.0000000000000000000000000000000000000001147735species00063.00000063.00000000000000000000000000000000000000011111263832strain00063.00000063.00000000000000000000000000000000000000025514016991196861111111116111128211class66.747.276.076.668.070.563.617.0076.8070.371.059.0069.068.00065.059.0069.00065.059.00000059.00000000071.059.0069.068.00032111111111111111356order61.044.5069.068.063.50000063.565.00069.068.00065.00069.00065.000000000000000065.00069.068.00021331141294family044.50068.055.30000055.3000068.0000000000000000000000000000068.000212211374genus044.50068.042.50000042.5000068.0000000000000000000000000000068.00011111183627species000068.00000000000068.0000000000000000000000000000068.0002222631580no\_rank044.500042.50000042.50000000000000000000000000000000000001111288000species0000047.00000047.00000000000000000000000000000000000002112111325120species044.500038.00000038.0000000000000000000000000000000000000111073genus0000081.00000081.0000000000000000000000000000000000000111076species0000081.00000081.00000000000000000000000000000000000001111316055strain0000081.00000081.000000000000000000000000000000000000023345401family59.0000060.70000060.700000000000000000000000000000000000023381genus59.0000060.70000060.700000000000000000000000000000000000022253399species59.0000067.50000067.5000000000000000000000000000000000000222222670307strain59.0000067.50000067.5000000000000000000000000000000000000111427356species0000047.00000047.000000000000000000000000000000000000011111029756strain0000047.00000047.00000000000000000000000000000000000004469277family0000068.50000068.5000000000000000000000000000000000000441168287genus0000068.50000068.50000000000000000000000000000000000001171433species0000075.00000075.000000000000000000000000000000000000011111082933strain0000075.00000075.000000000000000000000000000000000000022325217no\_rank0000062.00000062.000000000000000000000000000000000000011112483404species0000044.00000044.000000000000000000000000000000000000011112654249species0000080.00000080.000000000000000000000000000000000000011111182115family65.00000000000065.000000065.00000065.000000000000000065.00000001111227290no\_rank65.00000000000065.000000065.000000000000000000000065.000000011111379genus65.00000000000065.000000065.000000000000000000000065.00000001112613769no\_rank65.00000000000065.0000000000000000000000000000065.00000001111112048897species65.00000000000065.0000000000000000000000000000065.0000000111111119045family00069.0077.00000077.000069.000000069.000000000000000000000069.000011407genus0000077.00000077.0000000000000000000000000000000000000112615210no\_rank0000077.00000077.000000000000000000000000000000000000011112603276species0000077.00000077.000000000000000000000000000000000000011111186650genus00069.00000000000069.000000069.000000000000000000000069.00001112617746no\_rank00069.00000000000069.0000000000000000000000000000069.00001111112082949species00069.00000000000069.0000000000000000000000000000069.000011766order7.0000007.00000000000000000000000000000000000000000011942family7.0000007.00000000000000000000000000000000000000000011952tribe7.0000007.00000000000000000000000000000000000000000011953genus7.0000007.000000000000000000000000000000000000000000112640676no\_rank7.0000007.0000000000000000000000000000000000000000001111163164species7.0000007.0000000000000000000000000000000000000000001111111204441order059.00000000000059.000000059.00000059.00000059.000000000059.000000111111433family059.00000000000059.000000059.00000059.000000000000000059.00000011111125216genus059.00000000000059.000000059.000000000000000000000059.0000001112617492no\_rank059.00000000000059.0000000000000000000000000000059.0000001111112018065species059.00000000000059.0000000000000000000000000000059.00000011313204455order69.071.000074.369.0000074.30000000000000000000000000000000000001131331989family69.071.000074.369.0000074.300000000000000000000000000000000000012211265genus071.000083.50000083.50000000000000000000000000000000000001111111545044species071.000075.00000075.000000000000000000000000000000000000011111060genus69.0000056.069.0000056.0000000000000000000000000000000000000111063species0000056.00000056.00000000000000000000000000000000000001111992186strain0000056.00000056.000000000000000000000000000000000000011111075species69.00000069.0000000000000000000000000000000000000000001711395371195233204457order69.817.076.076.8071.870.917.0076.8071.765.3000000000000000000000000000065.3000000171139537119523341297family69.817.076.076.8071.870.917.0076.8071.765.3000000000000000000000000000065.300000041394211941331113687genus69.517.0076.8073.5017.0076.8073.365.3000000000000000000000000000065.30000002139361193522196159no\_rank63.017.0076.8075.9017.0076.8075.863.0000000000000000000000000000063.000000011111327635species017.00000017.0000000000000000000000000000000000000000022221517554species0000079.50000079.500000000000000000000000000000000000039331932393319321523415species00076.8075.800076.8075.70000000000000000000000000000000000001111111938607species65.00000000000065.0000000000000000000000000000065.00000001111112565555species61.00000000000061.0000000000000000000000000000061.000000011112599297species0000070.00000070.0000000000000000000000000000000000000111111363835species70.00000000000070.0000000000000000000000000000070.000000011111560345species0000081.00000081.00000000000000000000000000000000000001331332071607species82.0000053.00000053.000000000000000000000000000000000000011112594473species0000051.00000051.000000000000000000000000000000000000051242165695genus66.2076.00070.073.0000070.0000000000000000000000000000000000000111113690species0000062.00000062.0000000000000000000000000000000000000511412611147no\_rank66.2076.00078.073.0000078.00000000000000000000000000000000000001111627192species0000078.00000078.00000000000000000000000000000000000005145142072936species66.2076.000073.0000000000000000000000000000000000000000008939111165696genus72.2000064.868.0000064.8000000000000000000000000000000000000121212121176536species56.0000039.056.0000039.000000000000000000000000000000000000066262644732no\_rank75.3000075.774.0000075.70000000000000000000000000000000000001111111609758species79.0000082.00000082.0000000000000000000000000000000000000552555252675225species74.6000074.474.0000074.400000000000000000000000000000000000022222204458order82.5000066.00000066.082.5000000000000000000000000000082.50000002222276892family82.5000066.00000066.082.5000000000000000000000000000082.50000002222241275genus82.5000066.00000066.082.5000000000000000000000000000082.5000000222222622653no\_rank82.5000066.00000066.082.5000000000000000000000000000082.50000002222221938605species82.50000000000082.5000000000000000000000000000082.500000022222562582species0000066.00000066.0000000000000000000000000000000000000124671965140416321040412241112111122428216class50.954.236.747.236.058.343.449.325.540.0058.373.056.09.021.800073.062.09.036.000062.007.0000007.000000073.056.09.021.8000111111132003order0007.0000000000007.00000007.0000007.0000007.00000000007.000011111190627family0007.0000000000007.00000007.0000007.00000000000000007.000011111453161genus0007.0000000000007.00000007.00000000000000000000007.00001112647919no\_rank0007.0000000000007.000000000000000000000000000007.00001111112559597species0007.0000000000007.000000000000000000000000000007.0000124641964139816321039812311112380840order50.953.836.747.836.058.343.449.325.540.0058.373.009.026.700073.009.065.000000000000000000073.009.026.700033506family0000011.70000011.700000000000000000000000000000000000011517genus0000020.00000020.00000000000000000000000000000000000001111519species0000020.00000020.000000000000000000000000000000000000022152267genus000007.5000007.500000000000000000000000000000000000022221940612species000007.5000007.5000000000000000000000000000000000000214421275682family009.0036.033.20000033.2009.00000009.00000000000000000000009.0000011221111963genus009.0036.039.50000039.5009.000000000000000000000000000009.000001111180842species009.0000000000009.000000000000000000000000000009.00000111078773strain009.0000000000000000000000000000000000000000000000111111863372species000036.037.00000037.00000000000000000000000000000000000001129580genus0000012.00000012.0000000000000000000000000000000000000112610881no\_rank0000012.00000012.000000000000000000000000000000000000011111644131species0000012.00000012.00000000000000000000000000000000000001111175654genus009.0000000000009.00000009.00000000000000000000009.000001112636909no\_rank009.0000000000009.000000000000000000000000000009.000001111112728020species009.0000000000009.000000000000000000000000000009.0000011202907genus0000042.00000042.000000000000000000000000000000000000011158899species0000042.00000042.000000000000000000000000000000000000011111005048strain0000042.00000042.00000000000000000000000000000000000001175917563781532103783131180864family50.754.539.948.1059.241.749.325.540.0059.200026.700000065.000000000000000000000026.700011111283genus00065.00000000000065.000000065.000000000000000000000065.00001111111562974species00065.00000000000065.0000000000000000000000000000065.00001731112916genus49.00032.70000023.3000006.000000000000000000000000000006.00002280867species0005.0000005.000000000000000000000000000000000000000221180870subspecies0005.0000005.0000000000000000000000000000000000000001111643561strain0005.0000005.0000000000000000000000000000000000000001111180869species0006.0000000000006.000000000000000000000000000006.000011397945strain0006.0000000000000000000000000000000000000000000001111553814species49.0006.000000000000000000000000000000000000000000000312684926no\_rank00069.00000060.00000000000000000000000000000000000000031312518343species00069.00000060.00000000000000000000000000000000000000011161634072genus0017.055.0043.20000043.20000000000000000000000000000000000001332234073species0017.00016.00000016.000000000000000000000000000000000000011595537strain0017.000000000000000000000000000000000000000000000011111246301strain0000033.00000033.00000000000000000000000000000000000009999436515species0000060.80000060.80000000000000000000000000000000000001441663243no\_rank00055.0024.20000024.200000000000000000000000000000000000033331795631species0000030.00000030.000000000000000000000000000000000000011112774875species000007.0000007.0000000000000000000000000000000000000116591345356153163561147420genus50.754.549.250.3060.841.749.344.042.2060.80009.000000000000000000000000000009.000011659134535615316356112610897no\_rank50.754.549.250.3060.841.749.344.042.2060.80009.000000000000000000000000000009.0000111111795665species0009.0000000000009.000000000000000000000000000009.000011659134435515316355116591344355153163552184519species50.754.549.251.2060.841.749.344.042.2060.800000000000000000000000000000000000011112565558species0000049.00000049.000000000000000000000000000000000000011180865genus0006.0013.00000013.000000000000000000000000000000000000011111180866species0006.0013.00000013.00000000000000000000000000000000000001111238749genus00077.006.000077.006.000000000000000000000000000000000000011112649760no\_rank00077.006.000077.006.000000000000000000000000000000000000011112714925species000006.0000006.000000000000000000000000000000000000011112735554species00077.00000077.0000000000000000000000000000000000000003313665874genus007.3009.3007.0009.300000000000000000000000000000000000033132626134no\_rank007.3009.3007.0009.3000000000000000000000000000000000000121212121678128species007.0009.0007.0009.00000000000000000000000000000000000002112111678129species007.50010.00000010.000000000000000000000000000000000000065813113119060family52.345.2045.8048.869.0000048.800000000000000000000000000000000000011148736genus69.087.0000069.000000000000000000000000000000000000000000111111329species69.087.0000069.00000000000000000000000000000000000000000033106589genus0000082.30000082.30000000000000000000000000000000000003333164546species0000082.30000082.300000000000000000000000000000000000054810101822464genus49.034.8045.8038.80000038.800000000000000000000000000000000000054810105481010134537species49.034.8045.8038.80000038.80000000000000000000000000000000000001111224471no\_rank73.00000000000073.000000073.000000000000000000000073.00000001111154066genus73.00000000000073.000000073.000000000000000000000073.00000001111112697032species73.00000000000073.0000000000000000000000000000073.00000003662112206351order062.700060.30000060.3056.000000062.00000062.000000000000000056.00000026611481family063.000060.30000060.3050.0000000000000000000000000000050.0000001111159genus050.000063.00000063.0050.0000000000000000000000000000050.00000011111163species050.00000000000050.0000000000000000000000000000050.000000112627922no\_rank0000063.00000063.0000000000000000000000000000000000000111196942species0000063.00000063.000000000000000000000000000000000000014411482genus076.000073.50000073.50000000000000000000000000000000000002222495species0000073.00000073.000000000000000000000000000000000000011111128449species076.000066.00000066.0000000000000000000000000000000000000111055692genus000005.0000005.00000000000000000000000000000000000001111682798species000005.0000005.00000000000000000000000000000000000001111111499392family062.00000000000062.000000062.00000062.000000000000000062.00000011111168470genus062.00000000000062.000000062.000000000000000000000062.000000111111168471species062.00000000000062.0000000000000000000000000000062.000000222122268525subphylum00048.50000000000048.500000048.50000047.00000048.50048.500000048.5000111111128221class00050.00000000000050.000000050.00000000000050.00050.000000050.000011111129order00050.00000000000050.000000050.00000000000050.000000000050.0000111180812suborder00050.00000000000050.000000050.000000000000000000000050.00001111215910no\_rank00050.00000000000050.000000050.000000000000000000000050.0000111111649470genus00050.00000000000050.000000050.000000000000000000000050.0000111111888845species00050.00000000000050.0000000000000000000000000000050.00001111111129547class00047.00000000000047.000000047.00000047.00000047.00047.000000047.00001111111213849order00047.00000000000047.000000047.00000047.00000047.000000000047.000011111172294family00047.00000000000047.000000047.00000047.000000000000000047.000011111194genus00047.00000000000047.000000047.000000000000000000000047.00001112593542no\_rank00047.00000000000047.0000000000000000000000000000047.00001111112561898species00047.00000000000047.0000000000000000000000000000047.000083532323no\_rank0009.4010.30009.0010.300000000000000000000000000000000000083531783234no\_rank0009.4010.30009.0010.3000000000000000000000000000000000000835395901no\_rank0009.4010.30009.0010.300000000000000000000000000000000000083532497643class0009.4010.30009.0010.300000000000000000000000000000000000083532497644order0009.4010.30009.0010.300000000000000000000000000000000000083532497645family0009.4010.30009.0010.300000000000000000000000000000000000083531551504genus0009.4010.30009.0010.300000000000000000000000000000000000083538353673862species0009.4010.30009.0010.30000000000000000000000000000000000001331132066phylum00053.0074.70000074.700053.0000000000000000000000000000053.000013311203490class00053.0074.70000074.700053.0000000000000000000000000000053.000013311203491order00053.0074.70000074.700053.0000000000000000000000000000053.0000133111129771family00053.0074.70000074.700053.0000000000000000000000000000053.00001331132067genus00053.0074.70000074.700053.0000000000000000000000000000053.00003340542species0000074.70000074.70000000000000000000000000000000000003333523794strain0000074.70000074.7000000000000000000000000000000000000111111157687species00053.00000000000053.0000000000000000000000000000053.000033340117phylum0009.0012.70000012.7000000000000000000000000000000000000333203693class0009.0012.70000012.7000000000000000000000000000000000000333189778order0009.0012.70000012.7000000000000000000000000000000000000333189779family0009.0012.70000012.7000000000000000000000000000000000000333179genus0009.0012.70000012.7000000000000000000000000000000000000333655606species\_group0009.0012.70000012.7000000000000000000000000000000000000333180species0009.0012.70000012.70000000000000000000000000000000000003333331162668strain0009.0012.70000012.700000000000000000000000000000000000012111157723phylum51.06.50006.051.06.00006.000000000000000000000000000000000000011204432class51.00000051.00000000000000000000000000000000000000000011204433order51.00000051.00000000000000000000000000000000000000000011204434family51.00000051.00000000000000000000000000000000000000000011392733genus51.00000051.0000000000000000000000000000000000000000001111392734species51.00000051.00000000000000000000000000000000000000000021111813735class06.50006.006.00006.000000000000000000000000000000000000021112211325family06.50006.006.00006.000000000000000000000000000000000000021112004797genus06.50006.006.00006.0000000000000000000000000000000000000211121111855912species06.50006.006.00006.00000000000000000000000000000000000007122101210111111111111111783257no\_rank60.027.07.036.0052.2007.036.0052.257.027.07.0000057.027.07.000057.027.07.0000027.07.0000000057.027.07.00000144111174201phylum57.0000045.80000045.857.000000057.00000057.000000000000000057.00000001441111203494class57.0000045.80000045.857.000000057.00000057.000000000000000057.0000000144111148461order57.0000045.80000045.857.000000057.00000057.000000000000000057.00000004411203557family0000045.80000045.800000000000000000000000000000000000022518753genus0000056.00000056.000000000000000000000000000000000000022222728835species0000056.00000056.0000000000000000000000000000000000000111348508genus0000039.00000039.0000000000000000000000000000000000000112711230no\_rank0000039.00000039.000000000000000000000000000000000000011112711231species0000039.00000039.00000000000000000000000000000000000001111111647988family57.00000000000057.000000057.00000057.000000000000000057.000000011111239934genus57.00000000000057.000000057.000000000000000000000057.0000000111111239935species57.00000000000057.0000000000000000000000000000057.000000061262611111203682phylum60.527.0036.0056.500036.0056.5027.000000027.00000027.00000027.000000000027.000000612626111113203683class60.527.0036.0056.500036.0056.5027.000000027.00000027.00000027.000000000027.0000001111111112order027.00000000000027.000000027.00000027.00000027.000000000027.000000111111126family027.00000000000027.000000027.00000027.000000000000000027.000000111112795774genus027.00000000000027.000000027.000000000000000000000027.0000001111112528007species027.00000000000027.0000000000000000000000000000027.000000326262691356order59.00036.0056.500036.0056.5000000000000000000000000000000000000326261763524family59.00036.0056.500036.0056.500000000000000000000000000000000000032626466152genus59.00036.0056.500036.0056.500000000000000000000000000000000000032626466153species59.00036.0056.500036.0056.50000000000000000000000000000000000003262632626886293strain59.00036.0056.500036.0056.50000000000000000000000000000000000002111111204428phylum007.0000007.0000007.00000007.0000007.0000007.00000000007.000002111111204429class007.0000007.0000007.00000007.0000007.0000007.00000000007.00000111111151291order007.0000000000007.00000007.0000007.0000007.00000000007.00000111111809family007.0000000000007.00000007.0000007.00000000000000007.0000011111113537no\_rank007.0000000000007.00000007.00000000000000000000007.0000011111810genus007.0000000000007.00000007.00000000000000000000007.0000011111813species007.0000000000007.000000000000000000000000000007.0000011759363strain007.0000000000000000000000000000000000000000000000111963360order007.0000007.00000000000000000000000000000000000000001192713family007.0000007.000000000000000000000000000000000000000011112987genus007.0000007.0000000000000000000000000000000000000000112643326no\_rank007.0000007.000000000000000000000000000000000000000011111353976species007.0000007.00000000000000000000000000000000000000002652044194313322213321783270no\_rank71.024.349.219.2062.2013.0016.6063.371.033.754.725.00000072.50000072.5000000000000071.033.754.725.000026520441943133222133268336no\_rank71.024.349.219.2062.2013.0016.6063.371.033.754.725.00000072.50000072.5000000000000071.033.754.725.0000265204419431332221332976phylum71.024.349.219.2062.2013.0016.6063.371.033.754.725.00000072.50000072.5000000000000071.033.754.725.00002618111810132132117743class71.024.3013.6023.6013.009.6024.471.033.7025.0000000000000000000000000071.033.7025.00002618111810132132200644order71.024.3013.6023.6013.009.6024.471.033.7025.0000000000000000000000000071.033.7025.0000261811181013213249546family71.024.3013.6023.6013.009.6024.471.033.7025.0000000000000000000000000071.033.7025.0000261891881321321111237genus71.024.3013.6015.2013.009.6015.171.033.7025.0000000000000000000000000071.033.7025.0000222222996species00025.00000000000025.0000000000000000000000000000025.00001111155197species014.00000000000014.0000000000000000000000000000014.000000111034807strain014.0000000000000000000000000000000000000000000000021315121214196345species016.0013.205.000011.005.000000000000000000000000000000000000011111452725strain00013.00000013.0000000000000000000000000000000000000002325124121211196869no\_rank71.033.306.5018.0013.006.5018.571.043.500000000000000000000000000071.043.5000002222935222species0006.5000006.50000000000000000000000000000000000000011111111111179672species71.013.000024.0013.000024.00000000000000000000000000000000000001111112249356species71.00000000000071.0000000000000000000000000000071.000000032322478552species0000018.70000020.00000000000000000000000000000000000001111112739062species07.0000000000007.000000000000000000000000000007.0000001111112748320species080.00000000000080.0000000000000000000000000000080.00000011312277species0000024.00000024.000000000000000000000000000000000000011111094466strain0000024.00000024.000000000000000000000000000000000000011111492737species0000011.00000011.00000000000000000000000000000000000001176831genus0000082.00000082.00000000000000000000000000000000000001111256species0000082.00000082.000000000000000000000000000000000000011111500genus0000041.00000041.00000000000000000000000000000000000001111516051species0000041.00000041.000000000000000000000000000000000000017711117747class0019.00075.60000075.60019.0000000000000000000000000000019.0000017711200666order0019.00075.60000075.60019.0000000000000000000000000000019.000001771184566family0019.00075.60000075.60019.0000000000000000000000000000019.000001111184567genus0019.00062.00000062.00019.0000000000000000000000000000019.00000111111363852species0019.00000000000019.0000000000000000000000000000019.00000112628915no\_rank0000062.00000062.000000000000000000000000000000000000011111727164species0000062.00000062.00000000000000000000000000000000000006611423349genus0000077.80000077.800000000000000000000000000000000000055423351species0000075.80000075.80000000000000000000000000000000000005555714943strain0000075.80000075.8000000000000000000000000000000000000210102222200643class0072.50080.00000080.00072.500000072.50000072.500000000000000072.50000210102222171549order0072.50080.00000080.00072.500000072.50000072.500000000000000072.5000011815family0000075.00000075.000000000000000000000000000000000000011816genus0000075.00000075.00000000000000000000000000000000000001111371601species0000075.00000075.0000000000000000000000000000000000000222222171552family0072.50000000000072.500000072.50000072.500000000000000072.5000022222838genus0072.50000000000072.500000072.500000000000000000000072.50000222222165179species0072.50000000000072.5000000000000000000000000000072.5000099333046no\_rank0000080.60000080.600000000000000000000000000000000000099909656genus0000080.60000080.600000000000000000000000000000000000066821species0000078.00000078.00000000000000000000000000000000000006666435590strain0000078.00000078.00000000000000000000000000000000000003333357276species0000085.70000085.70000000000000000000000000000000000002216116768503class0041.070.0071.900072.0071.90000000000000000000000000000000000002216116768507order0041.070.0071.900072.0071.9000000000000000000000000000000000000221611689373family0041.070.0071.900072.0071.90000000000000000000000000000000000002216116107genus0041.070.0071.900072.0071.9000000000000000000000000000000000000141414141379870species0000071.70000071.700000000000000000000000000000000000022212222122057025species0041.070.0073.000072.0073.000000000000000000000000000000000000014773022152931625191297464116072317592913532373372431441445241759445111783272no\_rank63.271.765.971.574.074.368.671.365.971.479.074.468.352.467.466.30077.671.665.081.061.0076.559.7000076.454.000074.772.0074.1074.068.252.467.466.30074.06815612961247527318243114531141239phylum59.070.559.754.049.070.172.269.756.755.3070.3068.563.80000065.081.00000000000000000000068.563.800005013975915652521515593339391061class63.173.275.659.3077.072.274.077.556.3077.1068.681.00000062.781.00000000000000000000068.681.00000501126471175221131166333631385order63.172.476.254.7076.672.273.983.053.4076.7064.081.00000062.781.00000000000000000000064.081.00000476194118931190964family81.576.878.00078.980.077.900079.0080.0000000000000000000000000000080.0000004761941189311186361279genus81.576.878.00078.980.077.900079.0080.0000000000000000000000000000080.00000032020220201280species063.000077.00000077.0000000000000000000000000000000000000111074919strain073.0000000000000000000000000000000000000000000000013426926134269261282species75.075.600076.8077.900076.8000000000000000000000000000000000000142114211283species84.074.800076.00000083.00000000000000000000000000000000000002131132131131290species081.000079.9082.000079.90000000000000000000000000000000000001111111292species080.00000000000080.0000000000000000000000000000080.0000001221101410122110141029380species80.081.278.00072.680.079.200072.60000000000000000000000000000000000001717151529388species0000084.60000084.6000000000000000000000000000000000000222272758subspecies0000082.00000082.00000000000000000000000000000000000002121283734species080.50000075.000000000000000000000000000000000000000004234247212411321535186817family60.262.168.054.7066.061.055.883.053.4066.0060.800000062.700000000000000000000060.800000423114717241317221386genus60.262.153.054.7064.161.055.8053.4064.1058.0000000000000000000000000000058.0000004224221397species075.000083.00000083.0000000000000000000000000000000000000221471species0000073.50000073.50000000000000000000000000000000000002222796606strain0000073.50000073.500000000000000000000000000000000000011111135841species055.00000000000055.0000000000000000000000000000055.0000004222147112213111186661species\_group60.259.453.054.7056.561.063.5053.4056.5061.0000000000000000000000000000061.000000111111392species061.00000000000061.0000000000000000000000000000061.000000111392837strain061.000000000000000000000000000000000000000000000004221147102213101396species60.259.353.054.7056.861.063.5053.4056.8000000000000000000000000000000000000422114710221310422114710221310526969strain60.259.353.054.7056.861.063.5053.4056.8000000000000000000000000000000000000111428species0000053.00000053.000000000000000000000000000000000000011180854no\_rank0000053.00000053.00000000000000000000000000000000000001111527030strain0000053.00000053.0000000000000000000000000000000000000111186664species028.00000028.000000000000000000000000000000000000000001111218284species068.00000068.00000000000000000000000000000000000000000211324767species083.500079.00000079.00000000000000000000000000000000000002112111367477strain083.500079.00000079.000000000000000000000000000000000000011653685species\_group0000077.00000077.000000000000000000000000000000000000011111402species0000077.00000077.00000000000000000000000000000000000001111145667genus045.00000000000045.000000045.000000000000000000000045.00000011111145668species045.00000000000045.0000000000000000000000000000045.00000011129337genus0000084.00000084.0000000000000000000000000000000000000112642459no\_rank0000084.00000084.000000000000000000000000000000000000011111233873species0000084.00000084.000000000000000000000000000000000000011150247genus0000072.00000072.0000000000000000000000000000000000000111133934species0000072.00000072.000000000000000000000000000000000000011111175304genus054.00000000000054.000000054.000000000000000000000054.0000001112614679no\_rank054.00000000000054.0000000000000000000000000000054.0000001111112518176species054.00000000000054.0000000000000000000000000000054.000000111112675230genus089.00000000000089.000000089.000000000000000000000089.00000011111665099species089.00000000000089.0000000000000000000000000000089.000000111196031strain089.0000000000000000000000000000000000000000000000012122800373genus0083.00070.50083.00070.5000000000000000000000000000000000000121212121404species0083.00070.50083.00070.5000000000000000000000000000000000000231333186818family73.0081.000077.0000000081.000000081.000000000000000000000081.00000333331372genus0081.00000000000081.000000081.000000000000000000000081.00000111111414778species0082.00000000000082.0000000000000000000000000000082.000001111111302659species0087.00000000000087.0000000000000000000000000000087.000001111111499687species0074.00000000000074.0000000000000000000000000000074.0000021160795genus73.00000077.000000000000000000000000000000000000000000212151173species73.00000077.00000000000000000000000000000000000000000011186822family0000086.00000086.00000000000000000000000000000000000001144249genus0000086.00000086.00000000000000000000000000000000000001111248903species0000086.00000086.000000000000000000000000000000000000022111539002no\_rank78.081.000079.082.0000079.0000000000000000000000000000000000000211539738no\_rank081.000079.00000079.00000000000000000000000000000000000002111378genus081.000079.00000079.00000000000000000000000000000000000002112111379species081.000079.00000079.000000000000000000000000000000000000021539742no\_rank78.00000082.0000000000000000000000000000000000000000002133986genus78.00000082.000000000000000000000000000000000000000000212644629no\_rank78.00000082.00000000000000000000000000000000000000000021211224749species78.00000082.00000000000000000000000000000000000000000027112393123933186826order076.672.077.4078.1075.072.075.0078.1077.7000000000000000000000000000077.70000027131331331300family076.6085.0078.2075.000078.2077.7000000000000000000000000000077.7000002713033033331301genus076.6085.0080.4075.000080.4077.7000000000000000000000000000077.7000003333331303species077.70000000000077.7000000000000000000000000000077.700000222332317233231308species076.500079.0075.000079.000000000000000000000000000000000000022322159strain076.00000000000000000000000000000000000000000000000331408178strain077.300000000000000000000000000000000000000000000001221111313species076.000077.50000077.50000000000000000000000000000000000001111574093strain0000088.00000088.0000000000000000000000000000000000000112211128037species077.0085.0083.00000083.0000000000000000000000000000000000000111111365659strain00085.0089.00000089.0000000000000000000000000000000000000111357genus0000013.00000013.000000000000000000000000000000000000011111363species0000013.00000013.0000000000000000000000000000000000000111133958family0072.00086.00072.00086.000000000000000000000000000000000000011111578genus0072.00086.00072.00086.00000000000000000000000000000000000001111111147770species0072.00086.00072.00086.000000000000000000000000000000000000011727186828family00076.7076.300075.0076.30000000000000000000000000000000000001172729393genus00076.7076.300075.0076.3000000000000000000000000000000000000117271172729394species00076.7076.300075.0076.300000000000000000000000000000000000018155361882138511186801class47.445.337.446.549.058.4016.015.050.7058.40012.0000000000000000000000000000012.000001168295order011.00000011.0000000000000000000000000000000000000000011543371family011.00000011.000000000000000000000000000000000000000001128895genus011.00000011.0000000000000000000000000000000000000000011111517species011.00000011.0000000000000000000000000000000000000000018145361881138511186802order47.447.737.446.549.058.4021.015.050.7058.40012.0000000000000000000000000000012.00000181453518112181131979family47.447.737.446.0046.2021.015.043.5046.20012.0000000000000000000000000000012.00000111266genus0000083.00000083.0000000000000000000000000000000000000112619470no\_rank0000083.00000083.000000000000000000000000000000000000011112726954species0000083.00000083.000000000000000000000000000000000000018145351711217111485genus47.447.737.446.0044.1021.015.043.5044.10012.0000000000000000000000000000012.0000018144351711217181443517112171513species47.447.743.846.0044.1021.015.043.5044.1000000000000000000000000000000000000111111169679species0012.00000000000012.0000000000000000000000000000012.00000535177186803family0000061.90000062.000000000000000000000000000000000000066841genus0000064.20000064.200000000000000000000000000000000000055166486species0000068.40000068.40000000000000000000000000000000000005555536231strain0000068.40000068.400000000000000000000000000000000000011301301species0000043.00000043.00000000000000000000000000000000000001111585394strain0000043.00000043.00000000000000000000000000000000000001128050genus0000035.00000035.00000000000000000000000000000000000001139485species0000035.00000035.00000000000000000000000000000000000001111515620strain0000035.00000035.000000000000000000000000000000000000011207244genus0000069.00000069.00000000000000000000000000000000000001111649756species0000069.00000069.00000000000000000000000000000000000002220572511genus0000060.20000060.3000000000000000000000000000000000000555533035species0000050.80000050.80000000000000000000000000000000000001191191912897species0000059.10000059.1000000000000000000000000000000000000662648079no\_rank0000070.00000070.000000000000000000000000000000000000066662479767species0000070.00000070.0000000000000000000000000000000000000771506553genus0000061.30000061.300000000000000000000000000000000000066661871021species0000062.30000062.3000000000000000000000000000000000000112608895no\_rank0000055.00000055.000000000000000000000000000000000000011111834196species0000055.00000055.0000000000000000000000000000000000000332316020genus0000068.00000068.00000000000000000000000000000000000003333038species0000068.00000068.00000000000000000000000000000000000003333411470strain0000068.00000068.0000000000000000000000000000000000000442569097genus0000070.00000070.0000000000000000000000000000000000000444439488species0000070.00000070.0000000000000000000000000000000000000112719231genus0000066.00000066.00000000000000000000000000000000000001184030species0000066.00000066.00000000000000000000000000000000000001111610130strain0000066.00000066.0000000000000000000000000000000000000112719313genus0000072.00000072.00000000000000000000000000000000000001111208479species0000072.00000072.00000000000000000000000000000000000001111186804family00065.0050.000065.0050.000000000000000000000000000000000000011111501226genus00065.0050.000065.0050.000000000000000000000000000000000000011111115758species0000050.00000050.0000000000000000000000000000000000000112626894no\_rank00065.00000065.00000000000000000000000000000000000000011112724150species00065.00000065.000000000000000000000000000000000000000111186806family000049.083.00000083.00000000000000000000000000000000000001111730genus000049.083.00000083.000000000000000000000000000000000000011129322species000049.083.00000083.0000000000000000000000000000000000000111111633697strain000049.083.00000083.000000000000000000000000000000000000033216572family0000070.00000070.000000000000000000000000000000000000033459786genus0000070.00000070.000000000000000000000000000000000000011351091species0000079.00000079.00000000000000000000000000000000000001111693746strain0000079.00000079.0000000000000000000000000000000000000222629304no\_rank0000065.50000065.500000000000000000000000000000000000022222763056species0000065.50000065.500000000000000000000000000000000000098541000family0000073.40000074.9000000000000000000000000000000000000321263genus0000056.30000053.5000000000000000000000000000000000000322608920no\_rank0000056.30000053.500000000000000000000000000000000000032322564099species0000056.30000053.500000000000000000000000000000000000066216851genus0000082.00000082.00000000000000000000000000000000000006666853species0000082.00000082.0000000000000000000000000000000000000332304686family000009.0000009.0000000000000000000000000000000000000332304692genus000009.0000009.0000000000000000000000000000000000000331515species000009.0000009.00000000000000000000000000000000000002222572545strain000009.0000009.000000000000000000000000000000000000011111138384strain000009.0000009.0000000000000000000000000000000000000111909932class00010.0012.00000012.0000000000000000000000000000000000000111909929order00010.0012.00000012.00000000000000000000000000000000000001111843490family00010.0012.00000012.0000000000000000000000000000000000000111365348genus00010.0012.00000012.0000000000000000000000000000000000000111365349species00010.0012.00000012.00000000000000000000000000000000000001111111192197strain00010.0012.00000012.00000000000000000000000000000000000002222221737404class068.500077.00000077.0068.500000068.500000000000000000000068.5000002222221737405order068.500077.00000077.0068.500000068.500000000000000000000068.5000002222221570339family068.500077.00000077.0068.500000068.500000000000000000000068.50000022150022genus0000077.00000077.000000000000000000000000000000000000022221260species0000077.00000077.000000000000000000000000000000000000022222162289genus068.50000000000068.500000068.500000000000000000000068.50000022222254005species068.50000000000068.5000000000000000000000000000068.5000001167819phylum0000037.00000037.0000000000000000000000000000000000000111077257class0000037.00000037.0000000000000000000000000000000000000111077263order0000037.00000037.0000000000000000000000000000000000000111077264family0000037.00000037.0000000000000000000000000000000000000111077265genus0000037.00000037.000000000000000000000000000000000000011454171species0000037.00000037.000000000000000000000000000000000000011111303518strain0000037.00000037.000000000000000000000000000000000000074573101417213641210144371135021619291223723724314414452261944514454201174phylum67.572.273.472.986.575.269.372.372.872.379.075.369.522.882.066.30077.671.70061.0076.554.0000076.454.000074.772.0074.1074.069.422.882.066.30074.059573101417213509101443711336206192911237137143441216194414413981760class68.572.273.472.986.575.473.372.372.872.379.075.569.322.882.066.30077.671.60061.0076.536.0000076.436.000074.70074.10069.322.882.066.30074.12526322624414611441610115416185006order76.262.365.071.3075.479.876.259.074.0075.476.17.882.076.200075.20065.000000000000000000075.57.882.076.200072132742266413174131268family73.161.3071.0078.7076.2074.0078.774.57.8065.000069.00065.000000000000000000073.67.8065.00003141141269genus075.7080.0075.5066.0080.0075.50000000000000000000000000000000000001111111270species00080.0080.000080.0080.00000000000000000000000000000000000001111465515strain00080.00000080.00000000000000000000000000000000000000033132620948no\_rank075.700074.0066.000074.0000000000000000000000000000000000000331333131179670species075.700074.0066.000074.0000000000000000000000000000000000000444444441663genus77.07.800073.80000073.877.07.800000000000000000000000000077.07.80000044444235627no\_rank77.0000073.80000073.877.0000000000000000000000000000077.0000000111111904039species73.00000000000073.0000000000000000000000000000073.00000003333331849032species78.30000000000078.3000000000000000000000000000078.300000044442600159species0000073.80000073.8000000000000000000000000000000000000444444656366species07.8000000000007.800000000000000000000000000007.8000008116211532207genus072.6068.0080.2082.0068.0080.30000000000000000000000000000000000008162158162152047species072.600080.2082.000080.30000000000000000000000000000000000001111169480species00068.00000068.0000000000000000000000000000000000000001111157493genus00065.00000000000065.000000065.000000000000000000000065.000011111172000species00065.00000000000065.0000000000000000000000000000065.000033131221742989genus076.700081.3075.000081.3000000000000000000000000000000000000212137929species081.50000075.00000000000000000000000000000000000000000112627139no\_rank0000065.00000065.000000000000000000000000000000000000011111155384species0000065.00000065.000000000000000000000000000000000000032331311742993genus68.00000000000069.500000069.000000000000000000000069.00000002222647000no\_rank69.50000000000069.5000000000000000000000000000069.50000002222222590775species69.50000000000069.5000000000000000000000000000069.500000031111185016family0065.00074.00059.00074.00082.0000000000000000000000000000082.000003111111707genus0065.00074.00059.00074.00082.0000000000000000000000000000082.0000021211708species0056.50000059.0000000000000000000000000000000000000000111112620175no\_rank0082.00074.00000074.00082.0000000000000000000000000000082.000001111112003551species0082.00000000000082.0000000000000000000000000000082.0000011112704467species0000074.00000074.000000000000000000000000000000000000054485020family066.200069.80000069.800000000000000000000000000000000000053343668genus066.200074.30000074.30000000000000000000000000000000000005335331331682species066.200074.30000074.300000000000000000000000000000000000011472568genus0000056.00000056.00000000000000000000000000000000000001111472569species0000056.00000056.00000000000000000000000000000000000006111112224485021family76.3000066.567.0000066.576.000000076.000000000000000000000076.000000011153357genus75.0000054.00000054.0000000000000000000000000000000000000111153358species75.0000054.00000054.00000000000000000000000000000000000001111710696strain0000054.00000054.0000000000000000000000000000000000000361699479genus77.0000063.867.0000063.800000000000000000000000000000000000036162643346no\_rank77.0000063.867.0000063.8000000000000000000000000000000000000361636161813880species77.0000063.867.0000063.800000000000000000000000000000000000011111367298genus76.00000000000076.000000076.000000000000000000000076.00000001112637926no\_rank76.00000000000076.0000000000000000000000000000076.00000001111112714941species76.00000000000076.0000000000000000000000000000076.0000000111112805645genus76.00000000000076.000000076.000000000000000000000076.0000000111111443156species76.00000000000076.0000000000000000000000000000076.000000011191932196556585023family78.50071.4077.284.00074.0077.277.70078.400078.600000000000000000000077.70078.400043231573genus79.5000077.786.0000077.7000000000000000000000000000000000000422228447species79.5000073.586.0000073.50000000000000000000000000000000000003212321233013subspecies79.0000073.591.0000073.5000000000000000000000000000000000000111133014subspecies81.00000081.000000000000000000000000000000000000000000112626594no\_rank0000086.00000086.000000000000000000000000000000000000011112768071species0000086.00000086.00000000000000000000000000000000000001962655112034genus00071.4073.300074.0073.300078.4000000000000000000000000000078.40001422035species00068.90000074.000000000000000000000000000000000000000142142138532no\_rank00068.90000074.000000000000000000000000000000000000000333369373species0000073.30000073.300000000000000000000000000000000000052255257496no\_rank00078.4077.50000077.500078.4000000000000000000000000000078.40005555551561023species00078.40000000000078.4000000000000000000000000000078.400011112070337species0000083.00000083.000000000000000000000000000000000000011112795488species0000072.00000072.000000000000000000000000000000000000021111133882genus76.5000070.080.0000070.073.0000000000000000000000000000073.00000001111370764species80.00000080.0000000000000000000000000000000000000000001111111072463species73.00000000000073.0000000000000000000000000000073.0000000112609290no\_rank0000070.00000070.000000000000000000000000000000000000011112782169species0000070.00000070.00000000000000000000000000000000000001169578genus0000067.00000067.0000000000000000000000000000000000000112649013no\_rank0000067.00000067.000000000000000000000000000000000000011111978566species0000067.00000067.00000000000000000000000000000000000001196492genus0000063.00000063.0000000000000000000000000000000000000112627005no\_rank0000063.00000063.000000000000000000000000000000000000011112596916species0000063.00000063.000000000000000000000000000000000000011111235888genus85.00000000000085.000000085.000000000000000000000085.00000001111112079791species85.00000000000085.0000000000000000000000000000085.000000077447237genus0000084.90000084.9000000000000000000000000000000000000772626248no\_rank0000084.90000084.900000000000000000000000000000000000077771446794species0000084.90000084.900000000000000000000000000000000000022222881616genus82.00000000000082.000000082.000000000000000000000082.00000002222618217no\_rank82.00000000000082.0000000000000000000000000000082.00000002222222735133species82.00000000000082.0000000000000000000000000000082.0000000222222680004genus72.00000000000072.000000072.000000000000000000000072.00000002222222419774species72.00000000000072.0000000000000000000000000000072.00000005463413831098802429108712112185007order54.872.374.073.0075.5072.073.572.3075.619.053.0042.0000000000000000000000000019.053.0042.0000446041382109080242810791211211653family52.272.374.073.0075.5072.073.572.2075.619.053.0042.0000000000000000000000000019.053.0042.000044604138210908024281079121121414758751716genus52.272.374.073.0075.5072.073.572.2075.619.053.0042.0000000000000000000000000019.053.0042.0000133331719species067.000064.30000064.3000000000000000000000000000000000000111089446strain067.00000000000000000000000000000000000000000000000111128028species0000073.00000073.00000000000000000000000000000000000003338288species0000072.30000072.30000000000000000000000000000000000003333585529strain0000072.30000072.300000000000000000000000000000000000011111138301species00042.0052.00000052.0000000000000000000000000000000000000253019261902530192619038304species76.076.2069.0075.700063.5075.9000000000000000000000000000000000000333339791species0000072.30000072.3000000000000000000000000000000000000111143765species0000060.00000060.0000000000000000000000000000000000000112605173885101126051738851043769species079.0073.4076.700072.7076.8000000000000000000000000000000000000222243770species0000057.00000057.000000000000000000000000000000000000011882321623311516111882321623311516143990species38.072.579.571.0076.0071.969.067.9076.000000000000000000000000000000000000011111165058species19.00000000000019.0000000000000000000000000000019.000000021112111146827species063.500060.0069.000060.0000000000000000000000000000000000000111111156978species056.00000000000056.0000000000000000000000000000056.00000021144143161879species081.059.067.0071.4075.000071.50000000000000000000000000000000000002114414321144143645127strain081.059.067.0071.4075.000071.50000000000000000000000000000000000002222161896species055.00000055.0000000000000000000000000000000000000000012121212161899species0000077.60000077.60000000000000000000000000000000000004433169292species0000066.00000066.00000000000000000000000000000000000001111548476strain0000069.00000069.00000000000000000000000000000000000001111187491species00083.00000083.00000000000000000000000000000000000000011111349751species00042.00000000000042.0000000000000000000000000000042.0000111224162strain00042.00000000000000000000000000000000000000000000037139133713913401472species00067.0062.900068.4062.90000000000000000000000000000000000001111111072256species050.00000000000050.0000000000000000000000000000050.000000252152435225215243521979527species072.3081.0072.8072.900072.800000000000000000000000000000000000011112080757species0000077.00000077.000000000000000000000000000000000000011112594913species0000075.00000075.000000000000000000000000000000000000011421122624378no\_rank073.078.064.0070.50078.077.0070.500000000000000000000000000000000000011112754725species0078.00000078.000000000000000000000000000000000000000014212142122763010species073.0064.0070.500077.0070.500000000000000000000000000000000000011185025family65.0000086.00000086.00000000000000000000000000000000000001111827genus65.0000086.00000086.00000000000000000000000000000000000001111828species65.0000086.00000086.0000000000000000000000000000000000000111051973strain65.00000000000000000000000000000000000000000000000011111443894strain0000086.00000086.0000000000000000000000000000000000000317172805586family077.0076.0075.100076.0075.1000000000000000000000000000000000000317171847725genus077.0076.0075.100076.0075.100000000000000000000000000000000000031717317171528099species077.0076.0075.100076.0075.10000000000000000000000000000000000002684312218551714118332291373743324343685009order65.174.381.072.486.575.268.273.185.079.079.075.462.30049.00077.600057.0076.50000076.4000074.70000062.30049.00074.75843102163117141162293737373731957family71.074.381.077.186.576.970.073.185.079.079.076.900000077.60000076.50000076.4000000000000000076.511129404genus74.0000066.00000066.000000000000000000000000000000000000011129405species74.0000066.00000066.00000000000000000000000000000000000001111111032480strain74.0000066.00000066.000000000000000000000000000000000000048431021621171411612937373781912216genus70.274.381.077.186.577.070.073.185.079.079.077.000000077.60000076.5000000000000000000000076.544631021361111411352929145192113114111229291747species70.273.781.077.186.578.270.071.385.079.079.078.300000077.6000000000000000000000000000077.64444267747strain0000084.50000084.500000000000000000000000000000000000011909952strain080.0000000000000000000000000000000000000000000000021211031709strain0079.50000085.000000000000000000000000000000000000000011111122995strain0000092.00000092.0000000000000000000000000000000000000111134454strain00085.00000000000000000000000000000000000000000000011111234380strain0000073.00000073.000000000000000000000000000000000000077771276648strain0000078.30000078.300000000000000000000000000000000000021010551734925subspecies66.0000079.30000079.30000000000000000000000000000000000002112111114967strain66.0000062.00000062.000000000000000000000000000000000000044441114969strain0000079.80000079.8000000000000000000000000000000000000111905725subspecies70.00000070.00000000000000000000000000000000000000000011111091045strain70.00000070.0000000000000000000000000000000000000000003826626382662633011species074.900070.3076.300070.30000000000000000000000000000000000002122242132132185015family63.70049.0062.967.8000063.962.30049.000000057.000000000000000000062.30049.0000151212201141839genus64.00041.0061.957.0000062.900041.0000000000000000000000000000041.0000122122433659species72.0000080.50000080.500000000000000000000000000000000000033332045452species0000065.00000065.00000000000000000000000000000000000003763762518371species69.0000060.70000063.80000000000000000000000000000000000003113112558918species67.0000037.00000037.00000000000000000000000000000000000001111112589074species00041.00000000000041.0000000000000000000000000000041.000048281112615069no\_rank64.0000060.257.0000060.200000000000000000000000000000000000032322483798species63.30000057.00000000000000000000000000000000000000000011112582905species0000070.00000070.000000000000000000000000000000000000011112592334species0000082.00000082.000000000000000000000000000000000000033332663857species0000046.30000046.300000000000000000000000000000000000022222714938species0000056.50000056.5000000000000000000000000000000000000311332040genus62.3000083.00000083.062.3000000000000000000000000000062.3000000311332633570no\_rank62.3000083.00000083.062.3000000000000000000000000000062.30000003333332079793species62.30000000000062.3000000000000000000000000000062.300000011112662028species0000083.00000083.00000000000000000000000000000000000002286795genus78.50000078.5000000000000000000000000000000000000000002222642780species78.50000078.50000000000000000000000000000000000000000011111116071genus00057.00000000000057.000000057.000000000000000000000057.000011111175385species00057.00000000000057.0000000000000000000000000000057.0000111111185011order36.00000000000036.000000036.00000036.00000036.000000000036.00000001111112062family36.00000000000036.000000036.00000036.000000000000000036.0000000111111883genus36.00000000000036.000000036.000000000000000000000036.000000011111183656species36.00000000000036.0000000000000000000000000000036.000000012285013order51.0000058.00000058.000000000000000000000000000000000000012274712family51.0000058.00000058.00000000000000000000000000000000000001221854genus51.0000058.00000058.00000000000000000000000000000000000001111859species51.0000058.00000058.0000000000000000000000000000000000000111111326424strain51.0000058.00000058.0000000000000000000000000000000000000112632575no\_rank0000058.00000058.00000000000000000000000000000000000001111710111species0000058.00000058.0000000000000000000000000000000000000221643682order0000075.50000075.50000000000000000000000000000000000002285030family0000075.50000075.50000000000000000000000000000000000002238501genus0000075.50000075.500000000000000000000000000000000000022138336species0000075.50000075.500000000000000000000000000000000000022221146883strain0000075.50000075.5000000000000000000000000000000000000111111643684order80.0000060.00000060.080.0000000000000000000000000000080.00000001111185031family80.0000060.00000060.080.0000000000000000000000000000080.00000001111153460genus80.0000060.00000060.080.0000000000000000000000000000080.00000001111153461species80.00000000000080.0000000000000000000000000000080.000000011479431strain80.00000000000000000000000000000000000000000000000011111090615species0000060.00000060.0000000000000000000000000000000000000121231284992class62.4000056.557.3000056.5000000000000000000000000000000000000121231284993order62.4000056.557.3000056.50000000000000000000000000000000000003111633392family66.3000040.075.0000040.000000000000000000000000000000000000031112789775genus66.3000040.075.0000040.0000000000000000000000000000000000000311131112789776species66.3000040.075.0000040.00000000000000000000000000000000000009112112448023family61.1000058.048.5000058.0000000000000000000000000000000000000911211682522genus61.1000058.048.5000058.0000000000000000000000000000000000000911211467094species61.1000058.048.5000058.00000000000000000000000000000000000009112119112111313172strain61.1000058.048.5000058.00000000000000000000000000000000000001111111184995class72.00000000000072.000000072.00000072.00000072.0000072.0000072.0000000111111184996order72.00000000000072.000000072.00000072.00000072.000000000072.00000001111112600303family72.00000000000072.000000072.00000072.000000000000000072.0000000111112600304genus72.00000000000072.000000072.000000000000000000000072.0000000111111496014species72.00000000000072.0000000000000000000000000000072.000000011184998class69.0000068.00000068.000000000000000000000000000000000000011184999order69.0000068.00000068.000000000000000000000000000000000000011184107family69.0000068.00000068.0000000000000000000000000000000000000111102106genus69.0000068.00000068.000000000000000000000000000000000000011174426species69.0000068.00000068.0000000000000000000000000000000000000111111411903strain69.0000068.00000068.0000000000000000000000000000000000000111497346class0000051.00000051.000000000000000000000000000000000000011588673order0000051.00000051.000000000000000000000000000000000000011320583family0000051.00000051.000000000000000000000000000000000000011191494genus0000051.00000051.000000000000000000000000000000000000011191495species0000051.00000051.00000000000000000000000000000000000001111469383strain0000051.00000051.00000000000000000000000000000000000005115132181321121798711no\_rank56.89.0045.4059.555.09.0057.1059.555.500000071.00000071.000000000000000055.500000051151321813211211117phylum56.89.0045.4059.555.09.0057.1059.555.500000071.00000071.000000000000000055.500000051221221121161order56.80060.0039.555.00060.0039.555.500000071.00000071.000000000000000055.5000000311111111162family59.30060.0070.075.00060.0070.040.0000000000000000000000000000040.0000000111163genus0000070.00000070.0000000000000000000000000000000000000112619674no\_rank0000070.00000070.000000000000000000000000000000000000011112490939species0000070.00000070.00000000000000000000000000000000000003111111177genus59.30060.00075.00060.00040.0000000000000000000000000000040.0000000111111306274species40.00000000000040.0000000000000000000000000000040.0000000112038116strain40.00000000000000000000000000000000000000000000000021112593658no\_rank69.00060.00075.00060.000000000000000000000000000000000000000211121112572090species69.00060.00075.00060.000000000000000000000000000000000000000111182family35.00000035.000000000000000000000000000000000000000000111203genus35.00000035.000000000000000000000000000000000000000000112618749no\_rank35.00000035.00000000000000000000000000000000000000000011111137095species35.00000035.000000000000000000000000000000000000000000111111119859family71.00000000000071.000000071.00000071.000000000000000071.000000011111111782genus71.00000000000071.000000071.000000000000000000000071.00000001112649714no\_rank71.00000000000071.0000000000000000000000000000071.00000001111112099387species71.00000000000071.0000000000000000000000000000071.0000000111892259family000009.0000009.000000000000000000000000000000000000011748770genus000009.0000009.0000000000000000000000000000000000000112622029no\_rank000009.0000009.000000000000000000000000000000000000011111914871species000009.0000009.0000000000000000000000000000000000000138781301283subclass00045.8065.200056.7065.2000000000000000000000000000000000000551118order0000064.80000064.8000000000000000000000000000000000000551890464family0000064.80000064.800000000000000000000000000000000000055102231genus0000064.80000064.8000000000000000000000000000000000000552623012no\_rank0000064.80000064.800000000000000000000000000000000000055551173026species0000064.80000064.8000000000000000000000000000000000000133731150order00045.8066.000056.7066.0000000000000000000000000000000000000621892252family00021.00000039.500000000000000000000000000000000000000511205genus00011.40000010.000000000000000000000000000000000000000511206species00011.40000010.0000000000000000000000000000000000000005151203124strain00011.40000010.0000000000000000000000000000000000000001154304genus00069.00000069.000000000000000000000000000000000000000111160species00069.00000069.0000000000000000000000000000000000000001111388467strain00069.00000069.00000000000000000000000000000000000000073531892254family00067.0066.000063.6066.000000000000000000000000000000000000073531158genus00067.0066.000063.6066.00000000000000000000000000000000000007353482564species00067.0066.000063.6066.000000000000000000000000000000000000073537353179408strain00067.0066.000063.6066.000000000000000000000000000000000000012121890424order09.000057.509.000057.5000000000000000000000000000000000000111890426family09.0000009.00000000000000000000000000000000000000000111129genus09.0000009.00000000000000000000000000000000000000000112626047no\_rank09.0000009.0000000000000000000000000000000000000000011111173263species09.0000009.00000000000000000000000000000000000000000221890428family0000057.50000057.5000000000000000000000000000000000000221142genus0000057.50000057.5000000000000000000000000000000000000222640012no\_rank0000057.50000057.500000000000000000000000000000000000011111147species0000023.00000023.000000000000000000000000000000000000011111148species0000092.00000092.0000000000000000000000000000000000000111890505order0000057.00000057.0000000000000000000000000000000000000111890528family0000057.00000057.00000000000000000000000000000000000001154298genus0000057.00000057.00000000000000000000000000000000000001154299species0000057.00000057.00000000000000000000000000000000000001111251229strain0000057.00000057.00000000000000000000000000000000000002582047455764618222443404158871778222222246803663322157superkingdom59.670.854.970.7070.360.472.042.869.0071.08.50000008.5000008.5000008.50000000008.500000030181811361918106231011197176692222228890phylum53.071.049.567.0070.439.572.437.066.4071.18.50000008.5000008.5000008.50000000008.50000003018181136191810623101119717669222222290931no\_rank53.071.049.567.0070.439.572.437.066.4071.18.50000008.5000008.5000008.50000000008.5000000301818113619181052310111971766822222183963class53.071.049.567.0070.439.572.437.066.4071.18.50000008.5000008.5000008.50000000008.500000022222221644055order8.5000000000008.50000008.5000008.5000008.50000000008.50000002222221963271family8.5000000000008.50000008.5000008.50000000000000008.5000000222221209988genus8.5000000000008.50000008.50000000000000000000008.50000002222221048396species8.5000000000008.500000000000000000000000000008.500000028181811361918105231011197176681644060order56.271.049.567.0070.439.572.437.066.4071.100000000000000000000000000000000000028181811361918105231011197176681644061family56.271.049.567.0070.439.572.437.066.4071.1000000000000000000000000000000000000281818113619181052310111971766829287genus56.271.049.567.0070.439.572.437.066.4071.100000000000000000000000000000000000028181811361918105231011197176682623058no\_rank56.271.049.567.0070.439.572.437.066.4071.1000000000000000000000000000000000000281818113619181052310111971766828181811361918105231011197176682055836species56.271.049.567.0070.439.572.437.066.4071.100000000000000000000000000000000000011224756class0000078.00000078.0000000000000000000000000000000000000112191order0000078.00000078.0000000000000000000000000000000000000112194family0000078.00000078.00000000000000000000000000000000000001145989genus0000078.00000078.00000000000000000000000000000000000001111118126species0000078.00000078.000000000000000000000000000000000000022822934293471134230380571111783275no\_rank60.469.156.770.6053.961.468.144.768.9054.2000000000000000000000000000000000000228229342934711342303805711128889phylum60.469.156.770.6053.961.468.144.768.9054.20000000000000000000000000000000000002282293429347113423038057111183924class60.469.156.770.6053.961.468.144.768.9054.200000000000000000000000000000000000022822934293471134230380571112281order60.469.156.770.6053.961.468.144.768.9054.20000000000000000000000000000000000002282293429347113423038057111118883family60.469.156.770.6053.961.468.144.768.9054.200000000000000000000000000000000000022822934293471134230380571112284genus60.469.156.770.6053.961.468.144.768.9054.200000000000000000000000000000000000022822934293471134230380571112641160no\_rank60.469.156.770.6053.961.468.144.768.9054.2000000000000000000000000000000000000228229342934711342303805711122822934293471134230380571112200888species60.469.156.770.6053.961.468.144.768.9054.200000000000000000000000000000000000031362811539288109030231494198287625268147123148028146298950612841111517621451341211517668615303419930318014623385244117992759superkingdom71.670.669.770.459.275.169.969.168.269.434.075.447.85.046.741.000061.55.037.839.20046.0040.036.50046.00036.00040.0046.0047.85.046.741.0000115555749636552452365432632123027class52.753.951.258.1061.337.237.537.549.2061.30000000000000000000000000000000000002918164873292351329589342order44.248.339.358.3062.341.0037.049.4062.30000000000000000000000000000000000002918164873292351329589343family44.248.339.358.3062.341.0037.049.4062.3000000000000000000000000000000000000291816487329235132955528genus44.248.339.358.3062.341.0037.049.4062.30000000000000000000000000000000000002918164873292351329291815483325235032555529species44.248.339.358.3062.341.0037.049.4062.30000000000000000000000000000000000001441414414905079strain0014.039.0034.000040.0034.00000000000000000000000000000000000004311389342211341111589350order53.448.656.747.2050.831.037.539.040.0050.800000000000000000000000000000000000029532514121142896family55.442.259.244.4048.414.037.539.00048.400000000000000000000000000000000000029532514121143030genus55.442.259.244.4048.414.037.539.00048.4000000000000000000000000000000000000295325141211429532514121142898species55.442.259.244.4048.414.037.539.00048.400000000000000000000000000000000000013654191119589351family48.954.044.250.8050.748.00040.0050.70000000000000000000000000000000000001365419111977924genus48.954.044.250.8050.748.00040.0050.70000000000000000000000000000000000001365419111913654191119464988species48.954.044.250.8050.748.00040.0050.7000000000000000000000000000000000000313440113533751089390314711772871043681356203448281352989216283591121761145342121761321538181171833154no\_rank71.670.769.770.562.975.270.069.268.269.4075.451.05.046.741.000077.05.037.839.2000040.036.50000036.00040.000051.05.046.741.0000240412201280353116524194731596436500217611453421217624119254716474751kingdom48.248.547.649.95.054.642.440.941.248.1054.751.05.046.741.000077.05.037.839.2000040.036.50000036.00040.000051.05.046.741.000088362110796725169611112252no\_rank48.544.448.046.0047.545.138.545.035.5047.5000000000000000000000000000000000000612710973731415374761phylum49.345.152.746.4047.243.042.047.035.5047.2000000000000000000000000000000000000612710973731415372683659no\_rank49.345.152.746.4047.243.042.047.035.5047.2000000000000000000000000000000000000612710973731415371111451435class49.345.152.746.4047.243.042.047.035.5047.200000000000000000000000000000000000023612134478order42.040.3028.8053.000037.5053.000000000000000000000000000000000000023612134479family42.040.3028.8053.000037.5053.00000000000000000000000000000000000002361214815genus42.040.3028.8053.000037.5053.0000000000000000000000000000000000000236121109760species42.040.3028.8053.000037.5053.0000000000000000000000000000000000000236121236121645134strain42.040.3028.8053.000037.5053.0000000000000000000000000000000000000582288232313932451442order49.748.054.949.9049.743.042.052.041.1049.70000000000000000000000000000000000005822882323139321142503no\_rank49.748.054.949.9049.743.042.052.041.1049.7000000000000000000000000000000000000582288232313932100474genus49.748.054.949.9049.743.042.052.041.1049.7000000000000000000000000000000000000582288232313932109871species49.748.054.949.9049.743.042.052.041.1049.7000000000000000000000000000000000000582288232313932582288232313932684364strain49.748.054.949.9049.743.042.052.041.1049.7000000000000000000000000000000000000111931432231171order41.08.032.026.4029.70032.021.8029.700000000000000000000000000000000000011193143286113family41.08.032.026.4029.70032.021.8029.700000000000000000000000000000000000011193143286114genus41.08.032.026.4029.70032.021.8029.700000000000000000000000000000000000011193143111931431806994species41.08.032.026.4029.70032.021.8029.7000000000000000000000000000000000000541822122116029phylum45.447.233.042.4051.800036.0051.8000000000000000000000000000000000000431551516032suborder46.055.033.038.2045.000036.0045.000000000000000000000000000000000000011127974family042.000034.00000034.000000000000000000000000000000000000011127977genus042.000034.00000034.000000000000000000000000000000000000011111140302species042.000034.00000034.0000000000000000000000000000000000000421441436734family46.061.533.036.8047.800036.0047.80000000000000000000000000000000000004214414322336033genus46.061.533.036.8047.800036.0047.8000000000000000000000000000000000000216035species00036.50000036.0000000000000000000000000000000000000002121284813isolate00036.50000036.000000000000000000000000000000000000000111158839species18.0033.00044.00000044.000000000000000000000000000000000000011111111876142strain18.0033.00044.00000044.00000000000000000000000000000000000001131616469895no\_rank43.024.0049.3054.20000054.2000000000000000000000000000000000000111010586132genus024.0059.0059.10000059.1000000000000000000000000000000000000111010586133species024.0059.0059.10000059.1000000000000000000000000000000000000111010111010881290strain024.0059.0059.10000059.100000000000000000000000000000000000012661633384genus43.00044.5046.00000046.0000000000000000000000000000000000000126612661485682species43.00044.5046.00000046.000000000000000000000000000000000000021510137411379135251913637phylum47.438.244.842.0045.246.835.037.00045.20000000000000000000000000000000000007341151115214504subphylum50.636.742.042.0043.235.0037.00043.20000000000000000000000000000000000007341151115214506class50.636.742.042.0043.235.0037.00043.2000000000000000000000000000000000000734115111536750order50.636.742.042.0043.235.0037.00043.2000000000000000000000000000000000000734115111536751family50.636.742.042.0043.235.0037.00043.200000000000000000000000000000000000073411511151129544genus50.636.742.042.0043.235.0037.00043.20000000000000000000000000000000000007341151115588596species50.636.742.042.0043.235.0037.00043.200000000000000000000000000000000000073411511157341151115747089strain50.636.742.042.0043.235.0037.00043.200000000000000000000000000000000000033451507subphylum0000035.00000035.0000000000000000000000000000000000000332212703class0000035.00000035.0000000000000000000000000000000000000334827order0000035.00000035.0000000000000000000000000000000000000331344966family0000035.00000035.0000000000000000000000000000000000000334836genus0000035.00000035.0000000000000000000000000000000000000334837species0000035.00000035.00000000000000000000000000000000000003333763407strain0000035.00000035.00000000000000000000000000000000000005131411141137986subphylum34.635.036.00044.344.035.000044.30000000000000000000000000000000000005131411142212732class34.635.036.00044.344.035.000044.3000000000000000000000000000000000000513141114214503order34.635.036.00044.344.035.000044.30000000000000000000000000000000000005131411144854family34.635.036.00044.344.035.000044.3000000000000000000000000000000000000513141114299330genus34.635.036.00044.344.035.000044.300000000000000000000000000000000000051314111451314111464571species34.635.036.00044.344.035.000044.30000000000000000000000000000000000002292117312503399163811877115362163572176114534212176324174652206582611932657451864subkingdom48.248.747.650.05.054.742.340.941.148.5054.851.05.046.741.000077.05.037.839.2000040.036.50000036.00040.000051.05.046.741.0000861400429112812397722352169239021661135242121661915533492217484890phylum47.246.744.143.45.053.841.140.732.436.0053.951.05.050.841.000077.05.043.039.2000049.036.50000036.00040.000051.05.050.841.00006616517875636147511451866subphylum39.540.843.838.7045.636.827.338.735.3045.60051.0000000000000000000000000000051.0000021111147553class30.5051.000029.0000000051.0000000000000000000000000000051.000002111137987order30.5051.000029.0000000051.0000000000000000000000000000051.000002111144281family30.5051.000029.0000000051.0000000000000000000000000000051.000002111114753genus30.5051.000029.0000000051.0000000000000000000000000000051.00000114754species29.00000029.00000000000000000000000000000000000000000011111408658strain29.00000029.00000000000000000000000000000000000000000011111263815species0051.00000000000051.0000000000000000000000000000051.00000111069680strain0051.000000000000000000000000000000000000000000000064165078755361475147554class39.840.843.638.7045.638.427.338.735.3045.60000000000000000000000000000000000006416507875536147534346order39.840.843.638.7045.638.427.338.735.3045.6000000000000000000000000000000000000641650787553614754894family39.840.843.638.7045.638.427.338.735.3045.60000000000000000000000000000000000006416507875536147531105254895genus39.840.843.638.7045.638.427.338.735.3045.6000000000000000000000000000000000000111713111311171311134896species39.018.069.044.6052.639.018.000052.600000000000000000000000000000000000024154037171247174897species39.742.344.243.4044.132.032.042.040.6044.100000000000000000000000000000000000024154037171247172415403717124717402676strain39.742.344.243.4044.132.032.042.040.6044.10000000000000000000000000000000000003682338325384899species39.4036.928.4042.140.3032.023.0042.1000000000000000000000000000000000000368233832538368233832538483514strain39.4036.928.4042.140.3032.023.0042.1000000000000000000000000000000000000122866546species0008.0057.00000057.0000000000000000000000000000000000000122122653667strain0008.0057.00000057.000000000000000000000000000000000000077636937310171227364184514822672156113524212156140332480253151211253716545no\_rank48.046.844.143.55.054.142.041.831.635.7054.251.05.050.841.000077.05.043.039.2000049.036.50000036.00040.000051.05.050.841.00002349077281303235746303211121147537subphylum47.245.344.230.7048.440.247.031.924.5048.40043.550.00000031.050.00000000000000000000043.550.000023490772813032357463032111214891class47.245.344.230.7048.440.247.031.924.5048.40043.550.00000031.050.00000000000000000000043.550.0000234907728130323574630321112120941319112194892order47.245.344.230.7048.440.247.031.924.5048.40043.550.00000031.050.00000000000000000000043.550.000039131026373215371111111359512154893family49.448.951.347.2050.244.353.563.051.0050.20031.050.00000031.050.00000000000000000000031.050.0000111114953genus0031.00000000000031.000000031.000000000000000000000031.000001111114956species0031.00000000000031.0000000000000000000000000000031.00000177310252112585310111033170genus50.746.155.748.7050.448.5063.059.0050.4000000000000000000000000000000000000636311333169species52.7055.752.0031.751.0063.00031.700000000000000000000000000000000000063631136363113284811strain52.7055.752.0031.751.0063.00031.70000000000000000000000000000000000003211212321121245286species52.043.5033.0052.90000052.9000000000000000000000000000000000000654424113604genus48.8054.657.5059.000054.0059.0000000000000000000000000000000000000653121113608species48.8054.657.7066.000054.0066.00000000000000000000000000000000000006531216531211071381strain48.8054.657.7066.000054.0066.00000000000000000000000000000000000001331071379species00057.0056.70000056.70000000000000000000000000000000000001331331071380strain00057.0056.70000056.700000000000000000000000000000000000011111300275genus00050.00000000000050.000000050.000000000000000000000050.000011111381046species00050.00000000000050.0000000000000000000000000000050.000011559295strain00050.000000000000000000000000000000000000000000000212313374469genus50.047.0044.0038.000049.0038.000000000000000000000000000000000000021231336033species50.047.0044.0038.000049.0038.0000000000000000000000000000000000000212313212313436907strain50.047.0044.0038.000049.0038.0000000000000000000000000000000000000111196389genus45.0042.0000000000000000000000000000000000000000000000111142260species45.0042.000000000000000000000000000000000000000000000014142381081531624131531127319family48.345.448.542.1048.941.340.536.534.5048.90056.0000000000000000000000000000056.000001211127320genus0035.030.0029.000029.0029.00000000000000000000000000000000000001211127322species0035.030.0029.000029.0029.000000000000000000000000000000000000012111280587variety0035.030.0029.000029.0029.00000000000000000000000000000000000001211112111869754strain0035.030.0029.000029.0029.0000000000000000000000000000000000000141423710615216241215211231136910genus48.345.448.942.3049.141.340.536.535.0049.10056.0000000000000000000000000000056.00000161319589151369136911species42.639.548.142.1047.541.430.040.336.3047.500000000000000000000000000000000000016131958915136911613195891513691306902strain42.639.548.142.1047.541.430.040.336.3047.50000000000000000000000000000000000001252916456011116601111540022no\_rank49.148.050.742.1051.641.351.025.033.7051.60056.0000000000000000000000000000056.00000125291445601111660125291445601111660498019species49.148.052.142.1051.641.351.025.033.7051.60000000000000000000000000000000000001111111231522species0056.00000000000056.0000000000000000000000000000056.000001471734353family62.00038.0048.700038.0048.7000000000000000000000000000000000000147174951genus62.00038.0048.700038.0048.7000000000000000000000000000000000000147174952species62.00038.0048.700038.0048.70000000000000000000000000000000000001471714717284591strain62.00038.0048.700038.0048.700000000000000000000000000000000000024151511148322248241407no\_rank39.138.530.013.9039.934.707.011.9039.90000000000000000000000000000000000001155317045genus0034.034.0055.20000055.20000000000000000000000000000000000001155317047species0034.034.0055.20000055.2000000000000000000000000000000000000115511551382522strain0034.034.0055.20000055.2000000000000000000000000000000000000241514110433222431910789genus39.138.529.713.7038.134.707.011.9038.100000000000000000000000000000000000024151411043322243241514110433222435481species39.138.529.713.7038.134.707.011.9038.100000000000000000000000000000000000028313410830family47.50030.9048.000035.0048.0000000000000000000000000000000000000721245787genus00030.7054.000035.0054.00000000000000000000000000000000000007212721245607species00030.7054.000035.0054.00000000000000000000000000000000000002111410829genus47.50032.0036.00000036.000000000000000000000000000000000000021112111796027species47.50032.0036.00000036.0000000000000000000000000000000000000434919766764family040.031.331.2048.600010.0048.6000000000000000000000000000000000000421111766728genus040.029.510.0065.000010.0065.00000000000000000000000000000000000004211114929species040.029.510.0065.000010.0065.0000000000000000000000000000000000000421111421111294746strain040.029.510.0065.000010.0065.000000000000000000000000000000000000013881535325no\_rank0035.038.3046.50000046.5000000000000000000000000000000000000138836913genus0035.038.3046.50000046.5000000000000000000000000000000000000138836914species0035.038.3046.50000046.500000000000000000000000000000000000013881388379508strain0035.038.3046.50000046.50000000000000000000000000000000000007777271271156497family45.450.147.634.4054.600011.0054.60000000000000000000000000000000000005674272713366genus50.249.547.643.0054.60000054.6000000000000000000000000000000000000567427275674272713502species50.249.547.643.0054.60000054.60000000000000000000000000000000000002131461281genus33.554.0023.00000011.00000000000000000000000000000000000000021312131460523species33.554.0023.00000011.000000000000000000000000000000000000000402246272656117172612369117112135112424212135111147538subphylum47.046.243.447.65.054.338.840.031.739.8054.451.05.055.739.200077.05.049.036.5000049.036.50000036.00040.000051.05.055.739.2000112155147549class36.044.055.042.0047.40000047.40000000000000000000000000000000000001121555185order36.044.055.042.0047.40000047.400000000000000000000000000000000000011215540289family36.044.055.042.0047.40000047.400000000000000000000000000000000000011215536048genus36.044.055.042.0047.40000047.400000000000000000000000000000000000011215539416species36.044.055.042.0047.40000047.4000000000000000000000000000000000000112155112155656061strain36.044.055.042.0047.40000047.400000000000000000000000000000000000011111111189478class00040.00000000000040.000000040.00000040.00000040.00040.000000040.00001111111189479order00040.00000000000040.000000040.00000040.00000040.000000000040.000011111147021family00040.00000000000040.000000040.00000040.000000000000000040.00001111113348genus00040.00000000000040.000000040.000000000000000000000040.00001111113349species00040.00000000000040.0000000000000000000000000000040.000011756982strain00040.00000000000000000000000000000000000000000000040124527065311711261236911705213411232312134601511141295134129716546no\_rank47.046.243.347.75.054.338.840.031.739.8054.451.05.055.739.000077.05.049.035.3000049.035.30000032.000000051.05.055.739.000013936409424671410246132213111147545class48.145.646.044.7052.043.444.038.534.3052.025.0055.700000049.00000049.0000000000000025.0055.7000022166451870subclass43.048.5039.0039.30000039.30000000000000000000000000000000000002216634395order43.048.5039.0039.30000039.30000000000000000000000000000000000002216643219family43.048.5039.0039.30000039.30000000000000000000000000000000000001555583genus00039.0039.00000039.00000000000000000000000000000000000001551033840species00039.0039.00000039.00000000000000000000000000000000000001551551182545strain00039.0039.00000039.0000000000000000000000000000000000000221143220genus43.048.500041.00000041.0000000000000000000000000000000000000221143228species43.048.500041.00000041.0000000000000000000000000000000000000221122111182542strain43.048.500041.00000041.00000000000000000000000000000000000001373440922397141023913221331466451871subclass48.245.546.044.7052.343.444.038.534.3052.325.0055.700000049.00000049.0000000000000025.0055.7000011927297821463821412221212136165042order49.444.946.544.9052.343.5038.735.5052.325.0049.000000049.00000049.0000000000000025.0049.0000021128568family35.0026.032.0000000000000000000000000000000000000000000002115094genus35.0026.032.0000000000000000000000000000000000000000000002112752537section35.0026.032.00000000000000000000000000000000000000000000021137727species35.0026.032.000000000000000000000000000000000000000000000211211441960strain35.0026.032.000000000000000000000000000000000000000000000116252574208628208111131492family49.745.446.544.8052.343.5026.535.5052.325.0000000000000000000000000000025.0000000116252574207628207112126224225052genus49.745.446.544.8052.443.5026.535.5052.425.0000000000000000000000000000025.00000001526131911941068species49.351.047.341.9048.10024.00048.100000000000000000000000000000000000015261319119152613191191392248strain49.351.047.341.9048.10024.00048.1000000000000000000000000000000000000517103481138151019species50.844.748.346.3053.742.00034.3053.7000000000000000000000000000000000000517103481138151710348113811448321strain50.844.748.346.3053.742.00034.3053.7000000000000000000000000000000000000111111209559species26.0000028.00000028.00000000000000000000000000000000000004444293939species0000046.20000046.20000000000000000000000000000000000008116381238319627species45.450.054.046.8054.846.00054.5054.8000000000000000000000000000000000000811638123881163812381450535strain45.450.054.046.8054.846.00054.5054.800000000000000000000000000000000000022411340412species43.5032.042.8078.00000078.000000000000000000000000000000000000022411224111392255strain43.5032.042.8078.00000078.000000000000000000000000000000000000011111487661species25.00000000000025.0000000000000000000000000000025.0000000111448322strain25.00000000000000000000000000000000000000000000000011111220188species0000050.00000050.000000000000000000000000000000000000013111340029species36.031.0011.00000011.000000000000000000000000000000000000000131113111448314strain36.031.0011.00000011.0000000000000000000000000000000000000001510493612362720871subgenus43.949.241.542.0052.60029.030.5052.60000000000000000000000000000000000002113445053species51.546.040.050.0041.50000041.5000000000000000000000000000000000000211344211344690307strain51.546.040.050.0041.50000041.500000000000000000000000000000000000013936321232306088species42.749.642.038.0054.00029.030.5054.000000000000000000000000000000000000013936321232139363212321392250strain42.749.642.038.0054.00029.030.5054.0000000000000000000000000000000000000121442720872subgenus44.0055.553.0044.00000044.0000000000000000000000000000000000000121225057species44.0055.553.0046.50000046.50000000000000000000000000000000000001212212122344612strain44.0055.553.0046.50000046.50000000000000000000000000000000000002236630species0000041.50000041.50000000000000000000000000000000000002222331117strain0000041.50000041.5000000000000000000000000000000000000115073genus0000028.00000028.000000000000000000000000000000000000011254878no\_rank0000028.00000028.0000000000000000000000000000000000000111108849species0000028.00000028.00000000000000000000000000000000000001111500485strain0000028.00000028.00000000000000000000000000000000000002222221131624family0049.00000000000049.000000049.00000049.000000000000000049.00000222225092genus0049.00000000000049.000000049.000000000000000000000049.00000222222264951species0049.00000000000049.0000000000000000000000000000049.000001571010191112191131111133183order40.547.645.242.5051.643.044.038.029.5051.60069.0000000000000000000000000000069.0000011111133184family46.044.0043.0056.0044.000056.000000000000000000000000000000000000011111133187genus46.044.0043.0056.0044.000056.000000000000000000000000000000000000011111133188species46.044.0043.0056.0044.000056.0000000000000000000000000000000000000111111111111336963strain46.044.0043.0056.0044.000056.000000000000000000000000000000000000051111134384family40.235.069.029.0000000000069.0000000000000000000000000000069.000005111115550genus40.235.069.029.0000000000069.0000000000000000000000000000069.0000051163400species40.235.0029.000000000000000000000000000000000000000000000511511663331strain40.235.0029.0000000000000000000000000000000000000000000001111163417species0069.00000000000069.0000000000000000000000000000069.0000011663202strain0069.000000000000000000000000000000000000000000000052527117299071family44.859.049.853.0053.343.0038.00053.3000000000000000000000000000000000000525261165036genus44.859.049.853.0061.043.0038.00061.0000000000000000000000000000000000000525261165037species44.859.049.853.0061.043.0038.00061.00000000000000000000000000000000000005252611652526116339724strain44.859.049.853.0061.043.0038.00061.000000000000000000000000000000000000011229219genus000007.0000007.0000000000000000000000000000000000000111681229species000007.0000007.00000000000000000000000000000000000001111559298strain000007.0000007.00000000000000000000000000000000000001245101101593277no\_rank14.046.033.540.8051.400016.0051.40000000000000000000000000000000000001115500genus14.051.057.00000000000000000000000000000000000000000000001115501species14.051.057.0000000000000000000000000000000000000000000000111111246410strain14.051.057.00000000000000000000000000000000000000000000001351011038946genus041.025.740.8051.400016.0051.400000000000000000000000000000000000032414121759species0025.725.5060.200016.0060.20000000000000000000000000000000000003241432414502780strain0025.725.5060.200016.0060.200000000000000000000000000000000000013661048829species041.0051.0045.50000045.500000000000000000000000000000000000013661366502779strain041.0051.0045.50000045.500000000000000000000000000000000000037141147547class48.30011.4044.00008.2044.0000000000000000000000000000000000000371411520881no\_rank48.30011.4044.00008.2044.000000000000000000000000000000000000037141388435subclass48.30011.4044.00008.2044.0000000000000000000000000000000000000371415197order48.30011.4044.00008.2044.000000000000000000000000000000000000037141157822suborder48.30011.4044.00008.2044.00000000000000000000000000000000000003714178060family48.30011.4044.00008.2044.000000000000000000000000000000000000037141112415genus48.30011.4044.00008.2044.00000000000000000000000000000000000003714137141112416species48.30011.4044.00008.2044.000000000000000000000000000000000000068578514514644114134611111111111715962no\_rank49.042.837.249.65.053.835.043.024.444.2054.077.05.0042.000077.05.0042.00000042.000000000000077.05.0042.000068578514514644114134611111111111111147541class49.042.837.249.65.053.835.043.024.444.2054.077.05.0042.000077.05.0042.00000042.000000000000077.05.0042.0000133159987no\_rank00025.0040.70000040.70000000000000000000000000000000000001111451869order00025.0030.00000030.00000000000000000000000000000000000001145131family0000030.00000030.00000000000000000000000000000000000001166739genus0000030.00000030.0000000000000000000000000000000000000111145133species0000030.00000030.000000000000000000000000000000000000022716585order0000046.00000046.000000000000000000000000000000000000022152637family0000046.00000046.000000000000000000000000000000000000022470095genus0000046.00000046.00000000000000000000000000000000000002222470096species0000046.00000046.000000000000000000000000000000000000012413351321111451867subclass0038.128.05.040.70030.85.0041.800042.000000042.00000042.000000000000000042.0000124133513211112726947order0038.128.05.040.70030.85.0041.800042.000000042.00000042.000000000000000042.000011111193133family00042.00000000000042.000000042.00000042.000000000000000042.0000111111047167genus00042.00000000000042.000000042.000000000000000000000042.0000111111047171species00042.00000000000042.0000000000000000000000000000042.000011336722strain00042.0000000000000000000000000000000000000000000001231335132668547family0038.123.35.040.70030.85.0041.800000000000000000000000000000000000022483074genus0000060.00000060.0000000000000000000000000000000000000221709381species0000060.00000060.00000000000000000000000000000000000002222717646strain0000060.00000060.000000000000000000000000000000000000012313151302072583genus0038.123.35.039.40030.85.0040.600000000000000000000000000000000000012313151301231315130245834species0038.123.35.039.40030.85.0040.600000000000000000000000000000000000068577313942741912425111111451868subclass49.042.837.050.5054.935.043.020.847.5055.077.05.00000077.05.00000000000000000000077.05.0000006857731394274191242511111192860order49.042.837.050.5054.935.043.020.847.5055.077.05.00000077.05.00000000000000000000077.05.00000033131717548648family60.042.330.031.7042.90000042.900000000000000000000000000000000000033131717548651genus60.042.330.031.7042.90000042.900000000000000000000000000000000000033131717548649species60.042.330.031.7042.90000042.900000000000000000000000000000000000033131717331317171392245strain60.042.330.031.7042.90000042.9000000000000000000000000000000000000655472136410419124081111111715340suborder48.542.837.150.9055.435.043.020.847.5055.577.05.00000077.05.00000000000000000000077.05.000000524459822982792965020family48.642.834.450.8056.136.5015.946.3056.3000000000000000000000000000000000000524459822982792961351751genus48.642.834.450.8056.136.5015.946.3056.30000000000000000000000000000000000005244598229827929613684species48.642.834.450.8056.136.5015.946.3056.30000000000000000000000000000000000005244598229827929652445982298279296321614strain48.642.834.450.8056.136.5015.946.3056.30000000000000000000000000000000000001111111128556family77.05.0000000000077.05.00000077.05.00000000000000000000077.05.000000111115598genus77.00000000000077.000000077.000000000000000000000077.00000001112499237section77.00000000000077.0000000000000000000000000000077.00000001111111187904species77.00000000000077.0000000000000000000000000000077.00000001111133194genus05.0000000000005.00000005.00000000000000000000005.000000111115016species05.0000000000005.000000000000000000000000000005.00000011665024strain05.00000000000000000000000000000000000000000000000651462634374family40.746.435.040.0042.733.5000042.700000000000000000000000000000000000065146265021genus40.746.435.040.0042.733.5000042.70000000000000000000000000000000000006514626220671no\_rank40.746.435.040.0042.733.5000042.700000000000000000000000000000000000065146265022species40.746.435.040.0042.733.5000042.70000000000000000000000000000000000006514626225342no\_rank40.746.435.040.0042.733.5000042.700000000000000000000000000000000000065146266514626985895strain40.746.435.040.0042.733.5000042.7000000000000000000000000000000000000641249106123106683158family50.248.550.851.8054.2043.038.051.0054.20000000000000000000000000000000000006412481011231015453genus50.248.550.852.0054.2043.038.051.0054.20000000000000000000000000000000000006412481011231016412481011231015454species50.248.550.852.0054.2043.038.051.0054.200000000000000000000000000000000000012255170genus00041.0045.00000045.0000000000000000000000000000000000000122100019species00041.0045.00000045.00000000000000000000000000000000000001221221150837strain00041.0045.00000045.000000000000000000000000000000000000033301206genus0000059.00000059.00000000000000000000000000000000000003333301207species0000059.00000059.00000000000000000000000000000000000001311371343938711091560868322131213323715989no\_rank44.547.546.048.4055.136.438.235.941.7055.200038.000000032.00000032.00000032.000000000038.000047114104316696281447694147548class40.748.046.349.4056.929.538.636.143.8056.90000000000000000000000000000000000002321092813128225178order39.338.033.741.4044.646.0026.08.0044.6000000000000000000000000000000000000555181family0000043.40000043.400000000000000000000000000000000000055101851genus0000043.40000043.400000000000000000000000000000000000055101852species0000043.40000043.400000000000000000000000000000000000055551116229strain0000043.40000043.4000000000000000000000000000000000000171961913191142128983family40.333.032.247.3045.146.0026.00045.100000000000000000000000000000000000036165179genus0033.70037.70021.00037.700000000000000000000000000000000000036165180species0033.70037.70021.00037.700000000000000000000000000000000000036163616665079isolate0033.70037.70021.00037.7000000000000000000000000000000000000162413111362111133196genus40.4025.049.0048.546.0027.00048.5000000000000000000000000000000000000104111110411111463999species40.70049.0049.60000049.600000000000000000000000000000000000011112478750species0000047.00000047.000000000000000000000000000000000000021176657family37.043.047.0000000000000000000000000000000000000000000000211324777genus37.043.047.0000000000000000000000000000000000000000000000211698440species37.043.047.0000000000000000000000000000000000000000000000211698441forma\_specialis37.043.047.00000000000000000000000000000000000000000000002112111072389strain37.043.047.0000000000000000000000000000000000000000000000112589077family37.00038.0000000000000000000000000000000000000000000001147830genus37.00038.00000000000000000000000000000000000000000000011111316788species37.00038.000000000000000000000000000000000000000000000322122755564family35.70025.5047.00008.0047.000000000000000000000000000000000000032212150173genus35.70025.5047.00008.0047.00000000000000000000000000000000000003221232212149040species35.70025.5047.00008.0047.00000000000000000000000000000000000002411294307668181146666221903no\_rank42.248.247.649.6057.413.038.638.844.5057.400000000000000000000000000000000000011106912946378104663534379family50.048.547.650.2057.9038.636.644.5058.000000000000000000000000000000000000011106912946378104663578156genus50.048.547.650.2057.9038.636.644.5058.0000000000000000000000000000000000000111069129463781046635111069129463781046635655981species50.048.547.650.2057.9038.636.644.5058.000000000000000000000000000000000000013631331113137240family35.541.747.335.4046.213.0061.00046.20000000000000000000000000000000000001363133111315100genus35.541.747.335.4046.213.0061.00046.20000000000000000000000000000000000001363133111315101species35.541.747.335.4046.213.0061.00046.2000000000000000000000000000000000000136313311131136313311131857342strain35.541.747.335.4046.213.0061.00046.200000000000000000000000000000000000072222777172611131713221371433147550class46.744.844.944.2048.238.735.033.034.2048.200038.000000032.00000032.00000032.000000000038.00005021226215031111149211295211191419222543subclass47.745.643.443.6047.436.035.033.033.2047.400041.000000032.00000032.000000000000000041.00001948326525651111115125order48.249.240.044.3049.238.50028.6049.200032.000000032.00000032.000000000000000032.00001111115129family00032.00000000000032.000000032.00000032.000000000000000032.0000111115543genus00032.00000000000032.000000032.000000000000000000000032.00001111129875species00032.00000000000032.0000000000000000000000000000032.000011413071strain00032.000000000000000000000000000000000000000000000193830652565110618family48.251.740.044.5049.238.50028.6049.2000000000000000000000000000000000000193830652565215506genus48.251.740.044.5049.238.50028.6049.2000000000000000000000000000000000000151313171631species\_group49.8014.00042.745.0000042.70000000000000000000000000000000000001513135507species49.8014.00042.745.0000042.700000000000000000000000000000000000015131359765forma\_specialis49.8014.00042.745.0000042.7000000000000000000000000000000000000151313151313426428strain49.8014.00042.745.0000042.700000000000000000000000000000000000043529621562569360species\_group42.251.741.444.7049.532.00028.6049.5000000000000000000000000000000000000435296215624352962156256646species42.251.741.444.7049.532.00028.6049.5000000000000000000000000000000000000225592order0000026.00000026.0000000000000000000000000000000000000225593family0000026.00000026.00000000000000000000000000000000000002241687genus0000026.00000026.00000000000000000000000000000000000002222563466species0000026.00000026.00000000000000000000000000000000000002212121964112631113441028384order46.438.845.643.7046.7035.033.027.0046.700050.0000000000000000000000000000050.0000117914231223681950family45.439.345.942.4048.20033.027.0048.200000000000000000000000000000000000011791423122312555455genus45.439.345.942.4048.20033.027.0048.200000000000000000000000000000000000012412707338no\_rank056.054.047.20000037.000000000000000000000000000000000000000124112411215731species056.054.047.20000037.000000000000000000000000000000000000000312112707348no\_rank45.329.054.517.00000017.0000000000000000000000000000000000000003121131870species45.329.054.517.00000017.0000000000000000000000000000000000000003121131211645133strain45.329.054.517.00000017.0000000000000000000000000000000000000008457181182707350no\_rank45.438.039.242.4047.30033.00047.300000000000000000000000000000000000084571811880884species45.438.039.242.4047.30033.00047.3000000000000000000000000000000000000845718118845718118759273strain45.438.039.242.4047.30033.00047.30000000000000000000000000000000000001023537136111033978family47.930.044.747.2045.4035.000045.400050.0000000000000000000000000000050.00001023537136111111036719genus47.930.044.747.2045.4035.000045.400050.0000000000000000000000000000050.00003112353427337species39.025.024.036.0045.50000045.50000000000000000000000000000000000003112353431123534498257strain39.025.024.036.0045.50000045.500000000000000000000000000000000000071211111051613species51.735.055.059.0036.0035.000036.000000000000000000000000000000000000071211117121111526221strain51.735.055.059.0036.0035.000036.00000000000000000000000000000000000001111111051616species00050.00000000000050.0000000000000000000000000000050.0000947161116111111313222544subclass35.6048.844.0053.147.00043.0053.100032.000000032.00000032.00000032.000000000032.0000436101102215139order36.0046.746.0049.200043.0049.2000000000000000000000000000000000000115148family0000035.00000035.0000000000000000000000000000000000000115140genus0000035.00000035.0000000000000000000000000000000000000115141species0000035.00000035.00000000000000000000000000000000000001111367110strain0000035.00000035.00000000000000000000000000000000000002349914435718family30.0046.750.0050.80000050.8000000000000000000000000000000000000233445149genus30.0046.745.3048.80000048.80000000000000000000000000000000000002334438033species30.0046.745.3048.80000048.80000000000000000000000000000000000002334423344306901strain30.0046.745.3048.80000048.8000000000000000000000000000000000000111920207genus0000034.00000034.00000000000000000000000000000000000001178579species0000034.00000034.00000000000000000000000000000000000001111573729strain0000034.00000034.00000000000000000000000000000000000001111111639021order00032.00000000000032.000000032.00000032.00000032.000000000032.000011111181093family00032.00000000000032.000000032.00000032.000000000000000032.00001111129849genus00032.00000000000032.000000032.000000000000000000000032.00001111136779species00032.00000000000032.0000000000000000000000000000032.000011644352strain00032.00000000000000000000000000000000000000000000041331775898order32.2055.00069.70000069.700000000000000000000000000000000000041331756146family32.2055.00069.70000069.7000000000000000000000000000000000000413365412genus32.2055.00069.70000069.70000000000000000000000000000000000004133223192species32.2055.00069.70000069.7000000000000000000000000000000000000413341331286976strain32.2055.00069.70000069.70000000000000000000000000000000000006143213222545subclass44.7061.045.8055.338.50036.0055.3000000000000000000000000000000000000614321337989order44.7061.045.8055.338.50036.0055.3000000000000000000000000000000000000111812776family0000050.00000050.00000000000000000000000000000000000001137840genus0000050.00000050.000000000000000000000000000000000000011393283species0000050.00000050.000000000000000000000000000000000000011111229662strain0000050.00000050.000000000000000000000000000000000000061422122033035family44.7061.045.8058.038.50036.0058.0000000000000000000000000000000000000614221242360genus44.7061.045.8058.038.50036.0058.000000000000000000000000000000000000061422126142212326645species44.7061.045.8058.038.50036.0058.00000000000000000000000000000000000001107599756205133268937924203310111149171314742214745204phylum48.349.649.753.8054.943.040.945.454.0054.90022.000000022.00000022.000000000000000022.000006403674959432155552455143215246173259421595302subphylum46.143.645.145.9052.841.431.840.842.3052.80000000000000000000000000000000000001566889249542131133054222155616class49.051.545.748.4054.841.247.039.749.9054.80000000000000000000000000000000000001566563246476131929476343135234order49.051.249.348.5056.241.247.042.250.7056.2000000000000000000000000000000000000376165215family45.3046.70039.30021.00039.300000000000000000000000000000000000037616105767genus45.3046.70039.30021.00039.3000000000000000000000000000000000000376165217species45.3046.70039.30021.00039.30000000000000000000000000000000000003761637616578456strain45.3046.70039.30021.00039.3000000000000000000000000000000000000153594824245513162845531884633family49.151.250.748.4056.541.247.040.250.6056.5000000000000000000000000000000000000149594124245513142845515206genus49.151.251.248.4056.541.247.044.850.6056.500000000000000000000000000000000000014957412394471314274471884637species\_group49.151.851.248.5056.441.247.044.850.9056.4000000000000000000000000000000000000149574123944713142744737769species49.151.851.248.5056.441.247.044.850.9056.400000000000000000000000000000000000014957412394471314274471495741239447131427447367775strain49.151.851.248.5056.441.247.044.850.9056.4000000000000000000000000000000000000138181897064species\_group07.0039.0061.600044.0061.6000000000000000000000000000000000000138185207species07.0039.0061.600044.0061.6000000000000000000000000000000000000311140410variety00039.0075.000044.0075.00000000000000000000000000000000000003131214684strain00039.00000044.0000000000000000000000000000000000000001111283643strain0000075.00000075.0000000000000000000000000000000000000177178876variety07.000059.70000059.7000000000000000000000000000000000000177177235443strain07.000059.70000059.7000000000000000000000000000000000000172490731genus45.0048.00000031.00000000000000000000000000000000000000001721721734106species45.0048.00000031.00000000000000000000000000000000000000003441211121910893family052.034.255.5059.80049.052.0059.80000000000000000000000000000000000003441211124998genus052.034.255.5059.80049.052.0059.80000000000000000000000000000000000003441211123441211124999species052.034.255.5059.80049.052.0059.800000000000000000000000000000000000032636441641851469order057.337.041.0044.50034.027.0044.500000000000000000000000000000000000032636441641759442family057.337.041.0044.50034.027.0044.500000000000000000000000000000000000048485552genus0000040.10000040.1000000000000000000000000000000000000484882508species0000040.10000040.10000000000000000000000000000000000004848189963variety0000040.10000040.1000000000000000000000000000000000000484848481186058strain0000040.10000040.10000000000000000000000000000000000003263164116105983genus057.337.041.0057.90034.027.0057.900000000000000000000000000000000000032631641163263164116105984species057.337.041.0057.90034.027.0057.900000000000000000000000000000000000043828240369215543821411131551631031323112155619class44.741.545.145.1052.140.731.041.040.3052.20000000000000000000000000000000000006336117210450531531450355688no\_rank44.043.150.649.1058.033.449.345.344.6058.0000000000000000000000000000000000000483077195391427313915303order44.143.251.949.4059.030.548.039.644.6059.0000000000000000000000000000000000000162115317family05.046.833.5000045.034.0000000000000000000000000000000000000001215324genus05.0033.50000034.0000000000000000000000000000000000000001215325species05.0033.50000034.000000000000000000000000000000000000000121121717944strain05.0033.50000034.00000000000000000000000000000000000000061114154genus0046.80000045.000000000000000000000000000000000000000061114155species0046.80000045.00000000000000000000000000000000000000006161732165strain0046.80000045.00000000000000000000000000000000000000004627681883874262938740465family44.744.752.749.8059.130.548.038.745.5059.10000000000000000000000000000000000004627681883874262938740466genus44.744.752.749.8059.130.548.038.745.5059.10000000000000000000000000000000000004627681883874262938746276818838742629387139825species44.744.752.749.8059.130.548.038.745.5059.100000000000000000000000000000000000021183979no\_rank00045.00000029.00000000000000000000000000000000000000021599838genus00045.00000029.0000000000000000000000000000000000000002121599839species00045.00000029.000000000000000000000000000000000000000223344396331family31.042.045.037.7047.50000047.50000000000000000000000000000000000002233445305genus31.042.045.037.7047.50000047.5000000000000000000000000000000000000223344231932species31.042.045.037.7047.50000047.5000000000000000000000000000000000000223344223344650164strain31.042.045.037.7047.50000047.500000000000000000000000000000000000061213212136064order51.253.053.046.2049.80000049.800000000000000000000000000000000000061213212157201family51.253.053.046.2049.80000049.800000000000000000000000000000000000061213212136065genus51.253.053.046.2049.80000049.80000000000000000000000000000000000006121321216121321211750568species51.253.053.046.2049.80000049.80000000000000000000000000000000000003327116139380order47.348.752.100045.052.053.3000000000000000000000000000000000000000332711640424family47.348.752.100045.052.053.30000000000000000000000000000000000000003327116167346genus47.348.752.100045.052.053.30000000000000000000000000000000000000003327116208960species47.348.752.100045.052.053.300000000000000000000000000000000000000033271163327116694068strain47.348.752.100045.052.053.300000000000000000000000000000000000000044452338order0000055.80000055.800000000000000000000000000000000000044908827family0000055.80000055.800000000000000000000000000000000000044133746genus0000055.80000055.800000000000000000000000000000000000044202698species0000055.80000055.80000000000000000000000000000000000004444741275strain0000055.80000055.80000000000000000000000000000000000006212452339order0039.80032.50031.00032.50000000000000000000000000000000000006212452340family0039.80032.50031.00032.5000000000000000000000000000000000000621240443genus0039.80032.50031.00032.50000000000000000000000000000000000006212104355species0039.80032.50031.00032.500000000000000000000000000000000000062126212670483strain0039.80032.50031.00032.5000000000000000000000000000000000000625232132452342order34.328.033.639.5052.90052.00052.90000000000000000000000000000000000001191940420family006.00056.20000056.20000000000000000000000000000000000001191913562genus006.00056.20000056.200000000000000000000000000000000000011919256003no\_rank006.00056.20000056.200000000000000000000000000000000000011919984962species006.00056.20000056.20000000000000000000000000000000000001191911919747525strain006.00056.20000056.2000000000000000000000000000000000000624213113103376family34.328.040.539.5048.10052.00048.10000000000000000000000000000000000006242131135644genus34.328.040.539.5048.10052.00048.100000000000000000000000000000000000062421311340492species34.328.040.539.5048.10052.00048.1000000000000000000000000000000000000624213113624213113721885strain34.328.040.539.5048.10052.00048.1000000000000000000000000000000000000369243276479109131182381108941143151715452333subclass44.841.442.643.3049.742.028.037.138.6049.7000000000000000000000000000000000000243154239305896231121508943813950143135145338order45.640.642.844.3050.142.724.337.638.5050.20000000000000000000000000000000000002357242245339family47.555.348.846.0049.00043.50049.00000000000000000000000000000000000002357242245340genus47.555.348.846.0049.00043.50049.00000000000000000000000000000000000002357242245341species47.555.348.846.0049.00043.50049.0000000000000000000000000000000000000235724224192523variety47.555.348.846.0049.00043.50049.0000000000000000000000000000000000000235724224235724224936046strain47.555.348.846.0049.00043.50049.00000000000000000000000000000000000001714539111395351family41.15.025.850.4048.431.005.049.0048.400000000000000000000000000000000000017145391113929882genus41.15.025.850.4048.431.005.049.0048.400000000000000000000000000000000000017145391113929883species41.15.025.850.4048.431.005.049.0048.4000000000000000000000000000000000000171453911139171453911139486041strain41.15.025.850.4048.431.005.049.0048.4000000000000000000000000000000000000941965108961272296104366family48.644.745.645.7056.644.6043.941.8056.60000000000000000000000000000000000009419651089612722965320genus48.644.745.645.7056.644.6043.941.8056.60000000000000000000000000000000000009419651089612722969419651089612722965322species48.644.745.645.7056.644.6043.941.8056.6000000000000000000000000000000000000921301261357237108227212024004family42.840.141.142.5049.341.624.631.134.8049.30000000000000000000000000000000000009213012613572371082272141247genus42.840.141.142.5049.341.624.631.134.8049.300000000000000000000000000000000000092130126135723710822721921301261357237108227212126181species42.840.141.142.5049.341.624.631.134.8049.30000000000000000000000000000000000008588371311807722418068889order42.142.641.840.8047.139.633.932.037.7047.1000000000000000000000000000000000000858837131180772241801227334suborder42.142.641.840.8047.139.633.932.037.7047.100000000000000000000000000000000000025101933453380634family48.736.5047.1052.943.20041.4052.900000000000000000000000000000000000025101933453380635genus48.736.5047.1052.943.20041.4052.900000000000000000000000000000000000025101933453380637species48.736.5047.1052.943.20041.4052.9000000000000000000000000000000000000251019334533251019334533741705strain48.736.5047.1052.943.20041.4052.900000000000000000000000000000000000060773711214737219147389951family39.443.441.839.7045.734.733.932.036.7045.70000000000000000000000000000000000006077371121473721914780744genus39.443.441.839.7045.734.733.932.036.7045.70000000000000000000000000000000000006077371121473721914785982species39.443.441.839.7045.734.733.932.036.7045.700000000000000000000000000000000000060773711214737219147341189variety39.443.441.839.7045.734.733.932.036.7045.70000000000000000000000000000000000006077371121473721914760773711214737219147578457strain39.443.441.839.7045.734.733.932.036.7045.70000000000000000000000000000000000004614156590231390111129000subphylum45.244.341.740.0051.645.0042.029.6051.60022.000000022.00000022.000000000000000022.0000096128251825162481class40.647.252.037.1054.059.00029.8054.000000000000000000000000000000000000096128251825231213order40.647.252.037.1054.059.00029.8054.0000000000000000000000000000000000000961282518251799696family40.647.252.037.1054.059.00029.8054.0000000000000000000000000000000000000961282518255533genus40.647.252.037.1054.059.00029.8054.00000000000000000000000000000000000009612825182529898species40.647.252.037.1054.059.00029.8054.00000000000000000000000000000000000009612825182596128251825578459strain40.647.252.037.1054.059.00029.8054.0000000000000000000000000000000000000378143765135651111162484class46.442.140.942.2050.731.0042.029.4050.70022.000000022.00000022.000000000000000022.000003781437651356511115258order46.442.140.942.2050.731.0042.029.4050.70022.000000022.00000022.000000000000000022.00000378133765135655259family46.442.142.442.2050.731.0042.029.4050.7000000000000000000000000000000000000378133765135655260genus46.442.142.442.2050.731.0042.029.4050.700000000000000000000000000000000000037813376513565203908species46.442.142.442.2050.731.0042.029.4050.70000000000000000000000000000000000003781337651356537813376513565747676strain46.442.142.442.2050.731.0042.029.4050.70000000000000000000000000000000000001111115262family0022.00000000000022.000000022.00000022.000000000000000022.00000111115296genus0022.00000000000022.000000022.000000000000000000000022.00000111115297species0022.00000000000022.0000000000000000000000000000022.00000156615forma\_specialis0022.000000000000000000000000000000000000000000000011418459strain0022.000000000000000000000000000000000000000000000037120123310299993011332609862121452284subphylum52.561.159.761.9059.345.759.552.961.6059.3000000000000000000000000000000000000418222754244545257class45.547.447.042.0051.740.0039.029.8051.7000000000000000000000000000000000000418222754244545267order45.547.447.042.0051.740.0039.029.8051.70000000000000000000000000000000000004182227542445411115268family45.547.447.042.0051.740.0039.029.8051.70000000000000000000000000000000000004082226522445263265genus45.447.447.041.8052.540.0039.029.8052.50000000000000000000000000000000000004082226522445240822265224452280036species45.447.447.041.8052.540.0039.029.8052.5000000000000000000000000000000000000111392992genus0000014.00000014.00000000000000000000000000000000000001184751species0000014.00000014.000000000000000000000000000000000000011111277687strain0000014.00000014.00000000000000000000000000000000000005116216452283class0042.633.0048.50041.00048.500000000000000000000000000000000000041161165404order0037.033.0048.50017.00048.500000000000000000000000000000000000011162920family00033.0023.00000023.0000000000000000000000000000000000000111215249genus00033.0023.00000023.0000000000000000000000000000000000000111111215250species00033.0023.00000023.0000000000000000000000000000000000000415115190068family0037.00050.20017.00050.2000000000000000000000000000000000000415115215251genus0037.00050.20017.00050.20000000000000000000000000000000000004151154151151280837species0037.00050.20017.00050.200000000000000000000000000000000000011162475order0065.00000065.000000000000000000000000000000000000000011162477no\_rank0065.00000065.0000000000000000000000000000000000000000111981958genus0065.00000065.000000000000000000000000000000000000000011111684307species0065.00000065.000000000000000000000000000000000000000032819220410009292811272569161538075class53.561.761.662.5060.046.159.555.962.1060.00000000000000000000000000000000000003281922041000929281127256916162474order53.561.761.662.5060.046.159.555.962.1060.00000000000000000000000000000000000003281922041000929281127256916742845family53.561.761.662.5060.046.159.555.962.1060.00000000000000000000000000000000000003281922041000929281127256916475941102662432655193genus53.561.761.662.5060.046.159.555.962.1060.0000000000000000000000000000000000000841184429454176773species55.6050.058.1055.845.90054.9056.800000000000000000000000000000000000084118442945418411844294541425265strain55.6050.058.1055.845.90054.9056.8000000000000000000000000000000000000174573280082711322078151745732800827113220781576775species52.057.864.363.7060.042.659.373.563.9059.90000000000000000000000000000000000001199715213576777species49.764.362.741.0059.2063.055.30059.200000000000000000000000000000000000011997152135119971521351230383strain49.764.362.741.0059.2063.055.30059.200000000000000000000000000000000000012673352924812912673352924812977020species55.464.255.854.0061.854.060.553.946.0061.80000000000000000000000000000000000001882204096subphylum50.0000050.00000050.0000000000000000000000000000000000000188431957class50.0000050.00000050.0000000000000000000000000000000000000188431958order50.0000050.00000050.0000000000000000000000000000000000000188431959family50.0000050.00000050.00000000000000000000000000000000000001881148959genus50.0000050.00000050.0000000000000000000000000000000000000881708541species0000050.00000050.00000000000000000000000000000000000008888671144strain0000050.00000050.0000000000000000000000000000000000000313198413521531088095314354862864501681161203375281192988566282939333208kingdom71.670.769.870.572.575.270.069.268.369.4075.500000000000000000000000000000000000031319841352153108809531435486286450168116120337528119298856628293936072no\_rank71.670.769.870.572.575.270.069.268.369.4075.5000000000000000000000000000000000000313198413521531088095314354862864501681161203375281192988566282939333213no\_rank71.670.769.870.572.575.270.069.268.369.4075.5000000000000000000000000000000000000313198413521531088095314354862864501681161203375281192988566282939333511no\_rank71.670.769.870.572.575.270.069.268.369.4075.500000000000000000000000000000000000031319841352153108809531435486286450168116120337528119298856628293937711phylum71.670.769.870.572.575.270.069.268.369.4075.5000000000000000000000000000000000000313198413521531088095314354862864501681161203375281192988566282939389593subphylum71.670.769.870.572.575.270.069.268.369.4075.500000000000000000000000000000000000031319841352153108809531435486286450168116120337528119298856628293937742no\_rank71.670.769.870.572.575.270.069.268.369.4075.500000000000000000000000000000000000031319841352153108809531435486286450168116120337528119298856628293937776no\_rank71.670.769.870.572.575.270.069.268.369.4075.50000000000000000000000000000000000003131984135215310880953143548628645016811612033752811929885662829393117570no\_rank71.670.769.870.572.575.270.069.268.369.4075.50000000000000000000000000000000000003131984135215310880953143548628645016811612033752811929885662829393117571no\_rank71.670.769.870.572.575.270.069.268.369.4075.500000000000000000000000000000000000031319841352153108809531435486286450168116120337528119298856628293938287superclass71.670.769.870.572.575.270.069.268.369.4075.500000000000000000000000000000000000031319841352153108809531435486286450168116120337528119298856628293931338369no\_rank71.670.769.870.572.575.270.069.268.369.4075.5000000000000000000000000000000000000313198413521531088095314354862864501681161203375281192988566282939332523no\_rank71.670.769.870.572.575.270.069.268.369.4075.5000000000000000000000000000000000000313198413521531088095314354862864501681161203375281192988566282939332524no\_rank71.670.769.870.572.575.270.069.268.369.4075.5000000000000000000000000000000000000313198413521531088095314354862864501681161203375281192988566282939340674class71.670.769.870.572.575.270.069.268.369.4075.5000000000000000000000000000000000000313198413521531088095314354862864501681161203375281192988566282939332525no\_rank71.670.769.870.572.575.270.069.268.369.4075.500000000000000000000000000000000000031319841352153108809531435486286450168116120337528119298856628293939347no\_rank71.670.769.870.572.575.270.069.268.369.4075.500000000000000000000000000000000000031319841352153108809531435486286450168116120337528119298856628293931437010no\_rank71.670.769.870.572.575.270.069.268.369.4075.50000000000000000000000000000000000003131984135215310880953143548628645016811612033752811929885662829393314146superorder71.670.769.870.572.575.270.069.268.369.4075.500000000000000000000000000000000000031319841352153108809531435486286450168116120337528119298856628293939443order71.670.769.870.572.575.270.069.268.369.4075.50000000000000000000000000000000000003131984135215310880953143548628645016811612033752811929885662829393376913suborder71.670.769.870.572.575.270.069.268.369.4075.50000000000000000000000000000000000003131984135215310880953143548628645016811612033752811929885662829393314293infraorder71.670.769.870.572.575.270.069.268.369.4075.500000000000000000000000000000000000031319841352153108809531435486286450168116120337528119298856628293939526parvorder71.670.769.870.572.575.270.069.268.369.4075.50000000000000000000000000000000000003131984135215310880953143548628645016811612033752811929885662829393314295superfamily71.670.769.870.572.575.270.069.268.369.4075.500000000000000000000000000000000000031319841352153108809531435486286450168116120337528119298856628293939604family71.670.769.870.572.575.270.069.268.369.4075.50000000000000000000000000000000000003131984135215310880953143548628645016811612033752811929885662829393207598subfamily71.670.769.870.572.575.270.069.268.369.4075.500000000000000000000000000000000000031319841352153108809531435486286450168116120337528119298856628293939605genus71.670.769.870.572.575.270.069.268.369.4075.5000000000000000000000000000000000000313198413521531088095314354862864501681161203375281192988566282939331319841352153108809531435486286450168116120337528119298856628293939606species71.670.769.870.572.575.270.069.268.369.4075.50000000000000000000000000000000000002161616554296no\_rank38.048.0047.7053.10000053.10000000000000000000000000000000000002161616172820family38.048.0047.7053.10000053.10000000000000000000000000000000000002161616877559genus38.048.0047.7053.10000053.10000000000000000000000000000000000002161616529818species38.048.0047.7053.10000053.100000000000000000000000000000000000021616162161616461836strain38.048.0047.7053.10000053.100000000000000000000000000000000000026151456249135249554915no\_rank41.149.143.144.8055.5051.039.733.8055.5000000000000000000000000000000000000261514562491352492605435phylum41.149.143.144.8055.5051.039.733.8055.500000000000000000000000000000000000032444142796class39.7042.525.5030.20000030.20000000000000000000000000000000000003244433083no\_rank39.7042.525.5030.20000030.2000000000000000000000000000000000000112058181order0044.029.000000000000000000000000000000000000000000000112058183family0044.029.00000000000000000000000000000000000000000000011133407genus0044.029.00000000000000000000000000000000000000000000011361139species0044.029.00000000000000000000000000000000000000000000011111410327strain0044.029.000000000000000000000000000000000000000000000313442058949order39.7041.024.3030.20000030.2000000000000000000000000000000000000313442058185family39.7041.024.3030.20000030.20000000000000000000000000000000000003134415782genus39.7041.024.3030.20000030.2000000000000000000000000000000000000112211225786species34.00013.0014.50000014.50000000000000000000000000000000000002112244689species42.5041.037.0046.00000046.00000000000000000000000000000000000002112221122352472strain42.5041.037.0046.00000046.000000000000000000000000000000000000023151252245135245555406no\_rank41.349.143.246.2055.9051.039.733.8055.9000000000000000000000000000000000000231512522451352452682482order41.349.143.246.2055.9051.039.733.8055.90000000000000000000000000000000000002315125224513524533084family41.349.143.246.2055.9051.039.733.8055.9000000000000000000000000000000000000231512522451352451335758genus41.349.143.246.2055.9051.039.733.8055.9000000000000000000000000000000000000282819411945759species52.053.0051.0059.100044.0059.10000000000000000000000000000000000002828194119428281941194294381strain52.053.0051.0059.100044.0059.100000000000000000000000000000000000011133085species41.000006.0000006.0000000000000000000000000000000000000111111370355strain41.000006.0000006.00000000000000000000000000000000000002071223471344746681species40.244.643.240.5044.8051.039.731.2044.80000000000000000000000000000000000002071223471344720712234713447370354strain40.244.643.240.5044.8051.039.731.2044.8000000000000000000000000000000000000702847143222435172222608109phylum48.141.646.646.5051.652.537.748.438.5051.6000000000000000000000000000000000000702847143222435172222830class48.141.646.646.5051.652.537.748.438.5051.6000000000000000000000000000000000000702847143222435172222608131no\_rank48.141.646.646.5051.652.537.748.438.5051.60000000000000000000000000000000000007028471432224351722273020order48.141.646.646.5051.652.537.748.438.5051.600000000000000000000000000000000000070284714322243517222418966family48.141.646.646.5051.652.537.748.438.5051.6000000000000000000000000000000000000702847143222435172222902genus48.141.646.646.5051.652.537.748.438.5051.6000000000000000000000000000000000000702847143222435172222903species48.141.646.646.5051.652.537.748.438.5051.60000000000000000000000000000000000007028471432224351722270284714322243517222280463strain48.141.646.646.5051.652.537.748.438.5051.600000000000000000000000000000000000011111112611341no\_rank46.00000000000046.000000046.00000046.00000046.0000000046.0046.000000011111111207245phylum46.00000000000046.000000046.00000046.00000046.0000000046.0046.000000011111115738order46.00000000000046.000000046.00000046.00000046.000000000046.00000001111115739family46.00000000000046.000000046.00000046.000000000000000046.0000000111168459subfamily46.00000000000046.000000046.000000000000000000000046.0000000111115740genus46.00000000000046.000000046.000000000000000000000046.00000001111115741species46.00000000000046.0000000000000000000000000000046.000000027510143011230112611352no\_rank47.452.251.546.0045.850.0045.047.0045.840.0000000000000000000000000000040.000000011555752phylum038.0043.0040.40000040.400000000000000000000000000000000000011552601529no\_rank038.0043.0040.40000040.400000000000000000000000000000000000011552601530no\_rank038.0043.0040.40000040.400000000000000000000000000000000000011555765family038.0043.0040.40000040.400000000000000000000000000000000000011555761genus038.0043.0040.40000040.400000000000000000000000000000000000011555762species038.0043.0040.40000040.400000000000000000000000000000000000011551155744533strain038.0043.0040.40000040.4000000000000000000000000000000000000274101325112251133682phylum47.455.851.546.2046.950.0045.047.0046.940.0000000000000000000000000000040.000000027410132511225115653class47.455.851.546.2046.950.0045.047.0046.940.0000000000000000000000000000040.000000027410132511225112704647subclass47.455.851.546.2046.950.0045.047.0046.940.0000000000000000000000000000040.000000027410132511225112704949order47.455.851.546.2046.950.0045.047.0046.940.0000000000000000000000000000040.000000027410132511225111225654family47.455.851.546.2046.950.0045.047.0046.940.0000000000000000000000000000040.0000000235321112115690genus47.7050.443.0040.550.0045.050.0040.50000000000000000000000000000000000001139700subgenus0000047.00000047.0000000000000000000000000000000000000115691species0000047.00000047.0000000000000000000000000000000000000115702subspecies0000047.00000047.00000000000000000000000000000000000001111185431strain0000047.00000047.0000000000000000000000000000000000000235311147570subgenus47.7050.443.00050.0045.050.00000000000000000000000000000000000000023531115693species47.7050.443.00050.0045.050.00000000000000000000000000000000000000023531112353111353153strain47.7050.443.00050.0045.050.000000000000000000000000000000000000000445921121111286322subfamily46.055.852.647.1047.700044.0047.740.0000000000000000000000000000040.000000012211115658genus40.0000042.00000042.040.0000000000000000000000000000040.0000000111111138568subgenus40.0000040.00000040.040.0000000000000000000000000000040.000000011138581species\_group40.00000000000040.0000000000000000000000000000040.0000000111115664species40.00000000000040.0000000000000000000000000000040.000000011347515strain40.0000000000000000000000000000000000000000000000003459191195683genus48.055.852.647.1048.300044.0048.3000000000000000000000000000000000000345919119345919119157538species48.055.852.647.1048.300044.0048.300000000000000000000000000000000000095332775585128412526594641731731251911358124301112302698737no\_rank45.070.541.843.934.050.137.669.134.137.134.050.151.0000000000000000000000000000051.0000000537324912235251116129461828721115811155418501115033630no\_rank45.970.844.945.434.051.136.769.238.939.434.051.151.0000000000000000000000000000051.0000000390324301533407792346162050776114891129681216685794phylum45.970.845.445.6052.436.369.237.540.2052.451.0000000000000000000000000000051.00000002183236494208470174612112646811422676class47.970.946.347.0056.637.769.339.142.4056.751.0000000000000000000000000000051.00000001193234045131340114612419338115819order52.570.949.650.3059.841.969.343.844.9060.051.0000000000000000000000000000051.00000001193234045131340114612419338111639119family52.570.949.650.3059.841.969.343.844.9060.051.0000000000000000000000000000051.000000011932340451313401146124193381194419681685820genus52.570.949.650.3059.841.969.343.844.9060.051.0000000000000000000000000000051.000000032321919155418101subgenus68.063.059.764.0058.30000058.3000000000000000000000000000000000000335821species0000031.30000031.300000000000000000000000000000000000033335823strain0000031.30000031.30000000000000000000000000000000000002335825species65.0000065.30000065.300000000000000000000000000000000000023323331271subspecies65.0000065.30000065.300000000000000000000000000000000000021775860species063.058.00067.40000067.40000000000000000000000000000000000002177217754757subspecies063.058.00067.40000067.400000000000000000000000000000000000012111121115861species74.0060.547.0044.00000044.0000000000000000000000000000000000000613231119411186461218117418103subgenus50.270.943.446.7060.146.069.361.037.9060.400000000000000000000000000000000000031225827species0010.711.0014.50000014.5000000000000000000000000000000000000312231221120755strain0010.711.0014.50000014.50000000000000000000000000000000000002432302122282246114812432302122282246114815855species52.070.947.650.1065.343.069.3043.5065.700000000000000000000000000000000000012125858species076.0070.0000000000000000000000000000000000000000000003784163441143452288species49.145.155.541.3050.447.559.061.032.2050.400000000000000000000000000000000000037841634411434378416344114341237626strain49.145.155.541.3050.447.559.061.032.2050.400000000000000000000000000000000000044221564127539126418104subgenus52.460.150.952.6058.037.0038.052.0058.200000000000000000000000000000000000044201563126538125442015631265381255849species52.461.550.952.9058.137.0038.054.5058.3000000000000000000000000000000000000211112111185471species045.5032.0043.000032.0043.0000000000000000000000000000000000000214581811111418107subgenus52.564.062.247.4066.900025.0066.951.0000000000000000000000000000051.000000012115833species54.0056.50048.00000048.00000000000000000000000000000000000001211121136329isolate54.0056.50048.00000048.0000000000000000000000000000000000000111111647221species51.00000000000051.0000000000000000000000000000051.0000000115616115616880535species064.064.047.4066.800025.0066.800000000000000000000000000000000000099244977130677130278112135211355863order42.442.343.241.3048.230.0036.435.7048.200000000000000000000000000000000000017311521122127994family42.738.344.843.6046.026.0038.00046.0000000000000000000000000000000000000173115211221323335873genus42.738.344.843.6046.026.0038.00046.00000000000000000000000000000000000001025131135872species40.0045.543.6046.326.0000046.30000000000000000000000000000000000001025131131025131131537102strain40.0045.543.6046.326.0000046.300000000000000000000000000000000000043121431215874species43.5044.70042.00038.00042.000000000000000000000000000000000000012115875species034.045.50050.00000050.000000000000000000000000000000000000012111211333668strain034.045.50050.00000050.000000000000000000000000000000000000013368886species0032.00040.70000040.7000000000000000000000000000000000000133133869250strain0032.00040.70000040.700000000000000000000000000000000000055132751743467432594family41.639.241.740.0048.926.7036.035.2048.9000000000000000000000000000000000000551327517434674269111115864genus41.639.241.740.0048.926.7036.035.2048.9000000000000000000000000000000000000231061924212245865species41.739.341.837.8049.529.0030.028.0049.50000000000000000000000000000000000002310619242122423106192421224484906no\_rank41.739.341.837.8049.529.0030.028.0049.5000000000000000000000000000000000000233323335866species42.5045.30047.00000047.00000000000000000000000000000000000001017133113315868species42.730.043.345.2049.80048.044.7049.80000000000000000000000000000000000001017133113311017133113311133968strain42.730.043.345.2049.80048.044.7049.80000000000000000000000000000000000001825103111318251031113189622species40.943.534.433.8047.722.0027.021.0047.700000000000000000000000000000000000022323723no\_rank0000049.50000049.50000000000000000000000000000000000002222462227species0000049.50000049.50000000000000000000000000000000000001245748103241528182401280412class43.543.144.744.1046.433.444.536.637.3046.4000000000000000000000000000000000000118544798228528172275796subclass44.143.645.144.6046.333.444.536.638.1046.40000000000000000000000000000000000001185447982285281722775739order44.143.645.144.6046.333.444.536.638.1046.400000000000000000000000000000000000011854479822852817227411388423054suborder44.143.645.144.6046.333.444.536.638.1046.400000000000000000000000000000000000036171232911136902236165799family41.943.239.241.8045.434.034.022.339.7045.40000000000000000000000000000000000003315122984113583611225800genus41.543.039.241.3044.034.034.022.338.6044.000000000000000000000000000000000000014221531143142215311435801species40.640.041.541.1039.034.034.0038.5039.0000000000000000000000000000000000000422471642247165802species42.545.046.541.2046.900039.0047.30000000000000000000000000000000000009888673679888673675804species41.044.636.940.6044.40022.30044.40000000000000000000000000000000000001144114444415species043.0047.0040.20000040.200000000000000000000000000000000000011111151315species051.000042.00000042.000000000000000000000000000000000000011144417genus37.0000047.00000047.000000000000000000000000000000000000011111188456species37.0000047.00000047.0000000000000000000000000000000000000573023397942679623135809family43.941.844.442.9044.733.2037.535.8044.70000000000000000000000000000000000004120232971325715810genus44.042.944.440.4044.732.3037.534.4044.70000000000000000000000000000000000004120232971325715811species44.042.944.440.4044.732.3037.534.4044.7000000000000000000000000000000000000412023297132571412023297132571508771strain44.042.944.440.4044.732.3037.534.4044.70000000000000000000000000000000000001010851594642genus44.939.6050.5044.000043.0044.0000000000000000000000000000000000000101085151010851594643species44.939.6050.5044.000043.0044.00000000000000000000000000000000000002161124501355035082family47.151.352.048.9050.3055.050.338.8050.300000000000000000000000000000000000021611245013550266265806genus47.151.352.048.9050.3055.050.338.8050.300000000000000000000000000000000000012225807species049.052.50042.50000042.500000000000000000000000000000000000012221222353152strain049.052.50042.50000042.500000000000000000000000000000000000012241633113335808species46.349.049.851.3046.9055.043.042.0046.900000000000000000000000000000000000012241633113331224163311333441375strain46.349.049.851.3046.9055.043.042.0046.900000000000000000000000000000000000052121237895species0053.649.5058.00054.00058.00000000000000000000000000000000000005212152121353151strain0053.649.5058.00054.00058.000000000000000000000000000000000000073887388857276species48.053.700055.20000055.200000000000000000000000000000000000063151311335086subclass31.333.027.034.2048.000025.0048.000000000000000000000000000000000000063151311335087order31.333.027.034.2048.000025.0048.000000000000000000000000000000000000063151311335088family31.333.027.034.2048.000025.0048.000000000000000000000000000000000000063151311335089genus31.333.027.034.2048.000025.0048.0000000000000000000000000000000000000631513113631513113110365species31.333.027.034.2048.000025.0048.000000000000000000000000000000000000022616132181325878phylum54.524.016.330.034.044.30010.032.834.044.30000000000000000000000000000000000002261613218132431838subphylum54.524.016.330.034.044.30010.032.834.044.300000000000000000000000000000000000022616132181326020class54.524.016.330.034.044.30010.032.834.044.3000000000000000000000000000000000000226161321813231277order54.524.016.330.034.044.30010.032.834.044.3000000000000000000000000000000000000226161311813137090suborder54.524.016.330.034.044.40010.032.834.044.400000000000000000000000000000000000022616131181315931genus54.524.016.330.034.044.40010.032.834.044.4000000000000000000000000000000000000226161311813122616131181315932species54.524.016.330.034.044.40010.032.834.044.40000000000000000000000000000000000001137093suborder0000042.00000042.000000000000000000000000000000000000011291294family0000042.00000042.0000000000000000000000000000000000000115890genus0000042.00000042.0000000000000000000000000000000000000115911species0000042.00000042.00000000000000000000000000000000000001111312017strain0000042.00000042.00000000000000000000000000000000000001305460151300616133002497438phylum45.546.745.946.5048.337.844.045.540.1048.300000000000000000000000000000000000013054601513006161330027997no\_rank45.546.745.946.5048.337.844.045.540.1048.300000000000000000000000000000000000013054601513006161330027998order45.546.745.946.5048.337.844.045.540.1048.300000000000000000000000000000000000013054601513006161330027999family45.546.745.946.5048.337.844.045.540.1048.300000000000000000000000000000000000013054601513006161330028000genus45.546.745.946.5048.337.844.045.540.1048.300000000000000000000000000000000000013054601513006161330031276species45.546.745.946.5048.337.844.045.540.1048.3000000000000000000000000000000000000130546015130061613300130546015130061613300423536strain45.546.745.946.5048.337.844.045.540.1048.300000000000000000000000000000000000037827636173513332922449913299541737123733634no\_rank43.542.239.842.7049.038.435.430.835.0049.1000000000000000000000000000000000000298194289538923221436709191114144762phylum42.439.438.441.7046.736.726.928.134.0046.800000000000000000000000000000000000014161664682329684763order34.934.537.637.8043.939.038.024.024.0043.90000000000000000000000000000000000001416166468232968741218112184764family34.934.537.637.8043.939.038.024.024.0043.900000000000000000000000000000000000071062840126404769genus32.035.141.338.0047.041.044.0017.0047.00000000000000000000000000000000000007106284012640101203species32.035.141.338.0047.041.044.0017.0047.000000000000000000000000000000000000071062840126407106284012640695850strain32.035.141.338.0047.041.044.0017.0047.000000000000000000000000000000000000021024102110100860genus041.535.335.7026.80024.035.0026.80000000000000000000000000000000000002102410211021024102110112090species041.535.335.7026.80024.035.0026.8000000000000000000000000000000000000283177273473851191134618474776order42.839.638.542.2046.937.023.928.335.4047.0000000000000000000000000000000000000283177273473851191134618474777family42.839.638.542.2046.937.023.928.335.4047.000000000000000000000000000000000000023234313314780genus54.013.048.349.8063.20055.70063.2000000000000000000000000000000000000232343133123234313314781species54.013.048.349.8063.20055.70063.200000000000000000000000000000000000028117425046982019113161816214783genus42.840.137.642.1046.337.023.925.635.4046.400000000000000000000000000000000000028117024346581619829608124787species42.840.737.742.2046.337.028.225.335.8046.400000000000000000000000000000000000028117024346581619829608122811702434658161982960812403677strain42.840.737.742.2046.337.028.225.335.8046.4000000000000000000000000000000000000214792species016.5000008.000000000000000000000000000000000000000002121761204strain016.5000008.00000000000000000000000000000000000000000254421142544211467593species014.525.228.8043.5014.58.015.0043.5000000000000000000000000000000000000113335675class30.00026.0024.70000024.7000000000000000000000000000000000000113354409order30.00026.0024.70000024.7000000000000000000000000000000000000113344055genus30.00026.0024.70000024.70000000000000000000000000000000000001133113344056species30.00026.0024.70000024.7000000000000000000000000000000000000334136115182445161822683628class44.245.444.346.1055.139.042.541.435.6055.1000000000000000000000000000000000000334136115182445161822683629no\_rank44.245.444.346.1055.139.042.541.435.6055.10000000000000000000000000000000000003341361151824451618242740order44.245.444.346.1055.139.042.541.435.6055.1000000000000000000000000000000000000334136115182445161822547934family44.245.444.346.1055.139.042.541.435.6055.10000000000000000000000000000000000003341361151824451618211412967genus44.245.444.346.1055.139.042.541.435.6055.1000000000000000000000000000000000000333733100179444121793337331001794441217912968species44.244.845.846.3055.439.042.545.036.0055.4000000000000000000000000000000000000434313944171no\_rank051.228.333.0037.30027.00037.3000000000000000000000000000000000000434313434313944170species051.228.333.0037.30027.00037.30000000000000000000000000000000000003736326418833311188612122696291no\_rank49.651.946.140.2051.750.058.345.739.5051.7000000000000000000000000000000000000242928471333331013362921122836phylum55.555.645.942.0053.450.058.345.742.6053.40000000000000000000000000000000000004421918251833836class51.864.035.042.6045.9071.5035.0045.90000000000000000000000000000000000004421918251833846subclass51.864.035.042.6045.9071.5035.0045.90000000000000000000000000000000000004421918251833847order51.864.035.042.6045.9071.5035.0045.90000000000000000000000000000000000004421918251829202family51.864.035.042.6045.9071.5035.0045.90000000000000000000000000000000000004421918251835127genus51.864.035.042.6045.9071.5035.0045.90000000000000000000000000000000000004421918251835128species51.864.035.042.6045.9071.5035.0045.90000000000000000000000000000000000004421918251844219182518296543strain51.864.035.042.6045.9071.5035.0045.900000000000000000000000000000000000014232619113213411333849class49.154.846.837.5054.839.532.045.750.0054.800000000000000000000000000000000000014232619113213411333850no\_rank49.154.846.837.5054.839.532.045.750.0054.800000000000000000000000000000000000014232619113213411338748order49.154.846.837.5054.839.532.045.750.0054.800000000000000000000000000000000000014232619113213411338749family49.154.846.837.5054.839.532.045.750.0054.80000000000000000000000000000000000001423261911321341132849genus49.154.846.837.5054.839.532.045.750.0054.80000000000000000000000000000000000001423261911321341132850species49.154.846.837.5054.839.532.045.750.0054.8000000000000000000000000000000000000142326191132134113142326191132134113556484strain49.154.846.837.5054.839.532.045.750.0054.8000000000000000000000000000000000000137411431435747class38.836.647.531.4042.20008.0042.200000000000000000000000000000000000013741143143425074order38.836.647.531.4042.20008.0042.200000000000000000000000000000000000013741143143425072family38.836.647.531.4042.20008.0042.2000000000000000000000000000000000000137411431435748genus38.836.647.531.4042.20008.0042.20000000000000000000000000000000000001374114314372520species38.836.647.531.4042.20008.0042.200000000000000000000000000000000000013741143143137411431431093141strain38.836.647.531.4042.20008.0042.2000000000000000000000000000000000000322543769no\_rank34.0000063.50000063.5000000000000000000000000000000000000322136419phylum34.0000063.50000063.500000000000000000000000000000000000032229197class34.0000063.50000063.5000000000000000000000000000000000000322227085genus34.0000063.50000063.5000000000000000000000000000000000000322322227086species34.0000063.50000063.500000000000000000000000000000000000013561552614929622582531132907111110239superkingdom77.377.573.254.8077.177.777.780.280.7077.30000024.00000024.00000024.0000000000000000024.0011111110442family0000024.00000000000024.00000024.00000024.0000000000000000024.0011111558017genus0000024.00000000000024.00000024.0000000000000000000000024.0011111156947species0000024.00000000000024.0000000000000000000000000000024.00135615525948296125825311329072559587no\_rank77.377.575.354.5077.277.777.780.280.7077.3000000000000000000000000000000000000135615525948296125825311329072732396kingdom77.377.575.354.5077.277.777.780.280.7077.3000000000000000000000000000000000000123616171172497569phylum35.637.334.832.5033.217.0000033.2000000000000000000000000000000000000123616171172497571subphylum35.637.334.832.5033.217.0000033.2000000000000000000000000000000000000123616171172497576class35.637.334.832.5033.217.0000033.2000000000000000000000000000000000000123616171171980410order35.637.334.832.5033.217.0000033.2000000000000000000000000000000000000123616171171980416family35.637.334.832.5033.217.0000033.20000000000000000000000000000000000001236161711711572genus35.637.334.832.5033.217.0000033.20000000000000000000000000000000000001236161711712361617117159150species35.637.334.832.5033.217.0000033.2000000000000000000000000000000000000134415495332294425725311328902732408phylum77.677.679.865.5077.477.977.780.280.7077.6000000000000000000000000000000000000134415495332294425725311328902732506class77.677.679.865.5077.477.977.780.280.7077.60000000000000000000000000000000000001344154953322944257253113289076804order77.677.679.865.5077.477.977.780.280.7077.6000000000000000000000000000000000000134415495332294425725311328902499399suborder77.677.679.865.5077.477.977.780.280.7077.60000000000000000000000000000000000001344154953322944257253113289011118family77.677.679.865.5077.477.977.780.280.7077.6000000000000000000000000000000000000134415495332294425725311328902501931subfamily77.677.679.865.5077.477.977.780.280.7077.600000000000000000000000000000000000013441549533229442572531132890694002genus77.677.679.865.5077.477.977.780.280.7077.6000000000000000000000000000000000000134415495332294425725311328902509511subgenus77.677.679.865.5077.477.977.780.280.7077.600000000000000000000000000000000000013441549533229442572531132890694009species77.677.679.865.5077.477.977.780.280.7077.600000000000000000000000000000000000013441549533229442572531132890134415495332294425725311328902697049no\_rank77.677.679.865.5077.477.977.780.280.7077.6000000000000000000000000000000000000212731341no\_rank0013.068.000000000000000000000000000000000000000000000212731360no\_rank0013.068.000000000000000000000000000000000000000000000212731618phylum0013.068.000000000000000000000000000000000000000000000212731619class0013.068.0000000000000000000000000000000000000000000002128883order0013.068.0000000000000000000000000000000000000000000002110699family0013.068.000000000000000000000000000000000000000000000212560142genus0013.068.000000000000000000000000000000000000000000000212560650species0013.068.00000000000000000000000000000000000000000000021211969841no\_rank0013.068.0000000000000000000000000000000000000000000001695553307528083397921517942787854no\_rank50.249.371.352.3076.850.846.871.141.0077.000000000000000000000000000000000000016955533075280833979215179428384no\_rank50.249.371.352.3076.850.846.871.141.0077.000000000000000000000000000000000000016955533075280833979215179481077no\_rank50.249.371.352.3076.850.846.871.141.0077.000000000000000000000000000000000000016955533075280833979215179416955533075280833979215179432630species50.249.371.352.3076.850.846.871.141.0077.0000000000000000000000000000000000000
